# Multiple system atrophy is associated with brain somatic mutations in clonal hematopoiesis genes

**DOI:** 10.64898/2026.03.17.26346081

**Authors:** Ben Thompson, Dominic Horner, Caoimhe Morley, Emil Gustavsson, Zane Jaunmuktane, Christos Proukakis

**Affiliations:** Department of Clinical and Movement Neurosciences, UCL Queen Square Institute of Neurology, London, UK; UK Dementia Research Institute, University of Cambridge, Cambridge, UK; Department of Clinical Neurosciences, School of Clinical Medicine, University of Cambridge, Cambridge, UK

**Keywords:** Multiple system atrophy, synucleinopathies, somatic mutations, duplex sequencing, clonal hematopoiesis

## Abstract

**Background:** Multiple system atrophy (MSA) is a sporadic disorder characterized by alpha-synuclein inclusions. Somatic mutations may contribute to neurodegeneration, with somatic copy number variants in *SNCA* (alpha-synuclein) reported in MSA, and somatic mutations in cancer driver genes in Alzheimer’s disease.

**Objectives:** To investigate whether small somatic mutations (single nucleotide variants [SNVs] and indels) in *SNCA*, *KCTD7*, and 10 cancer driver genes, including seven involved in clonal hematopoiesis (CH), are present in MSA brain.

**Methods:** We applied duplex sequencing with unique molecular identifiers to DNA from cerebellum, cingulate cortex, and putamen of 20 MSA cases (10 olivopontocerebellar atrophy [OPCA], 10 striatonigral degeneration [SND]) and nine controls.

**Results:** We detected 34 somatic mutations, including 14 in CH genes. CH gene mutations were elevated in MSA compared to controls, reaching significance in the cortex (p=0.033), most prominently in SND (p=0.011), which also showed putaminal enrichment (p=0.034). Multi-regional CH mutations were only detected in MSA (six cases, all brain regions in three). There were 10 coding CH mutations, eight of which were in MSA, including six multi-regional. MSA CH mutations included events with oncogenic potential. Both coding CH mutations in controls were restricted to the cerebellum. No coding variants were detected in *SNCA*, and neither *SNCA* nor *KCTD7* showed excess somatic mutations.

**Conclusions:** Somatic CH mutations are enriched in MSA brain. Their multi-regional presence and predicted oncogenic potential suggest a functionally relevant proliferative process extending beyond peripheral hematopoiesis. Somatic SNVs and indels in *SNCA* and *KCTD7* do not appear to be common MSA drivers.

## INTRODUCTION

Multiple system atrophy (MSA) is a rapidly progressive neurodegenerative disorder of unknown etiology. It can present with parkinsonian, cerebellar, and autonomic features, and is characterized by alpha-synuclein inclusions in the CNS.(1–3) MSA is a sporadic disease, with heritability <7%.(4) Inherited genetic variants are therefore unlikely to play a major role, although an association with *KCTD7* was recently reported.(5) Somatic mutations are acquired during development or aging, leading to mosaicism. They may have a role in neurodegenerative disorders, including synucleinopathies.(6),(7) We have demonstrated somatic copy number variants (CNVs, extra copies) of the alpha-synuclein gene (*SNCA*) in MSA and Parkinson’s disease (PD).(8,9) In MSA, these CNVs are associated with inclusions in the same cell, and exhibit a predilection for regions differentially affected in MSA pathological subtypes: the cerebellum in olivopontocerebellar atrophy (OPCA), and putamen in striatonigral degeneration (SND),(10) recently confirmed specifically in oligodendrocytes.(11)

The α-synuclein filaments in MSA are distinct from those observed in PD and Lewy body dementia (LBD), but with some similarities to juvenile onset synucleinopathy caused by a seven amino acid insertion.(12–14) The unique structure in MSA could arise from a different amino acid sequence, introduced by a somatic single nucleotide variant (SNV) or insertion / deletion (indel) affecting the coding region of *SNCA*. Individual neurons and oligodendrocytes in controls harbor hundreds of somatic mutations that accumulate linearly with age, with SNVs accumulating faster in oligodendrocytes and indels faster in neurons.(15)

Detection of somatic mutations at low variant allele fraction (VAF) in brain tissue requires targeted high-coverage sequencing. We previously used such a strategy on *SNCA* and other PD genes, and did not detect somatic SNVs in 26 PD and three MSA cases, using DNA predominantly from substantia nigra.(16) In this study, we refine our previous unique molecular identifier (UMI)-based approach by incorporating duplex sequencing. We target the coding exons of *SNCA*, including novel exons reported in low abundance alternative transcripts,(17) alongside *KCTD7*, and ten cancer driver genes. These were preferentially mutated in Alzheimer’s disease (AD) microglia,(18) and include seven involved in clonal hematopoiesis (CH). CH is an age-related phenomenon in which hematopoietic stem cells acquire somatic mutations that confer a survival advantage leading to clonal expansion within the peripheral blood, and is associated with hematological malignancies and non-neoplastic disorders,(19) but can also exacerbate solid tumor prognosis by direct infiltration.(20) CH has recently been observed to be elevated in blood both in MSA(21) and PD.(22) We therefore apply deep targeted sequencing with error correction to examine *SNCA*, *KCTD7*, and ten cancer driver genes in MSA and control brains, and report an association of MSA with brain CH mutations.

## MATERIALS AND METHODS

### MSA and control brain samples

We analyzed postmortem frozen cerebellum, putamen, and cingulate cortex from pathologically confirmed MSA-OPCA predominant (n=10) and MSA-SND predominant (n=10) cases and unaffected controls (n=9) (Fig. 1A), sourced from the Queen Square Brain Bank for Neurological Disorders (QSBB). Demographics are shown in Supplementary Table 1. Samples are coded based on the number used in our recent study of *SNCA* CNVs, as several overlapped.(11) A total of 86 brain regions were processed; cingulate cortex was unavailable for one patient (OPCA-02). All donors or their next of kin provided informed consent for use of their brain in research, and the study was approved by NHS Health Research Authority Ethics Committee London-Central (REC reference 23/LO/0044).

**Figure 1.**
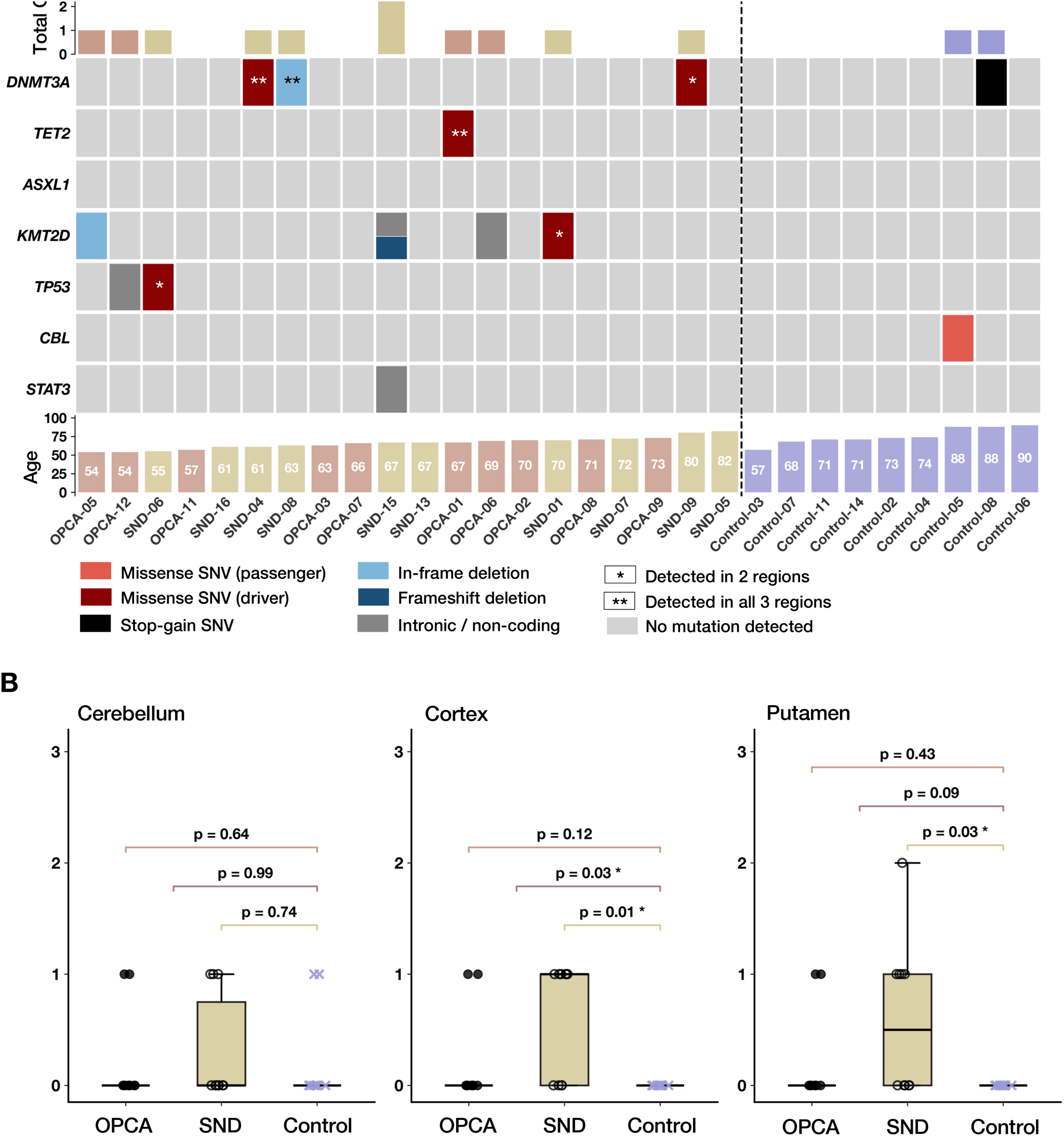
Study design and targeted duplex sequencing workflow. **A) Cohort overview:** post-mortem brain tissue from 10 MSA-OPCA, 10 MSA-SND, and 9 controls was sampled across three regions (cerebellum, cingulate cortex, putamen). Targeted NGS panel comprised MSA-associated genes and ten cancer driver genes; * denotes clonal hematopoiesis genes (B) UMI-based duplex sequencing: paired-end reads sharing UMI sequences are collapsed to generate consensus reads at three stringency levels (Duplex, Hybrid-MS2, Hybrid-MS1). **(C) Bioinformatic pipeline:** consensus BAMs processed through Mutect2 variant calling followed by candidate filtering and manual IGV inspection.

### Target gene panel and enrichment strategy

The genes selected were *SNCA*, *KCTD7*, and ten cancer driver genes recurrently mutated in AD brains (*BCR*, *MLH1*, *ATRX*, *ASXL1*, *CBL*, *DNMT3A*, *KMT2D*, *TET2*, *TP53*, *STAT3*), the last seven of which are involved in CH (Fig. 1A). Targeted capture was performed using SureSelect XT HS2 (Fig. 1B). This employs unique molecular identifiers (UMIs) to tag each DNA molecule before amplification, and minimises errors by generating consensus from reads sharing identical UMIs.(23) These are then used to generate duplex consensus sequences, removing any errors in one strand only.(24) This protocol has previously been successful in detecting low-frequency somatic mutations from frozen human brain tissue.(18) Probes were designed using the SureDesign portal, targeting coding exons and untranslated regions with an additional 10 bases from both ends. For *SNCA*, we additionally included enhancer and promoter regions, 15 exons recently identified as expressed at levels >1% of the total transcript,(17) and *SNCA-AS1*, a long noncoding antisense RNA overlapping the 5′ end that regulates *SNCA* expression and is associated with LBD.(25) The entire *SNCA* target is shown in Supplementary Fig. 1, and coordinates of all targets are summarized in Supplementary Table 2 (total size 168.5 kb).

### DNA library preparation and sequencing

DNA was extracted from 15–30 mg tissue using the QIAGEN MagAttract HMW DNA kit, followed by QC using TapeStation, NanoDrop, and Qubit. Libraries were generated from 100–200 ng DNA using the custom probe panel. Paired-end 150 bp sequencing was performed on Illumina NovaSeq X.

### Processing of sequencing data

FASTQ reads were trimmed using AGeNT Trim (v3.1.2) and aligned to GRCh38 with BWA-MEM (v0.7.18) using the -C flag to retain UMI tags. BAM files were sorted using samtools (v1.21) and underwent deduplication and error correction using AGeNT CReaK (v1.1.1). Error correction via read collapse produced consensus reads at three stringency levels: full duplex consensus; Hybrid-MS2, which includes single-strand consensus reads supported by a minimum of two reads; and Hybrid-MS1, which includes all UMI families without a minimum read requirement, maximizing sensitivity. Read group information was added to consensus BAMs using Picard (v3.3.0) AddOrReplaceReadGroups.

### Somatic mutation calling and filtering pipeline

To benchmark somatic mutation detection, we generated artificial mosaic mixtures using DNA from a patient brain with *SNCA* G51D mutation (26) into a control at dilutions of 1%, 0.5%, and 0.25%, producing VAFs of 0.5%, 0.25%, and 0.125% for heterozygous, and 1%, 0.5%, and 0.25% for homozygous SNPs. Germline SNPs unique to the G51D sample were called using HaplotypeCaller (GATK v4.6.1.0) on raw and duplex-filtered BAMs, and filtered using strand-bias (SOR ≤ 3), mapping-quality (MQ ≥ 40), read-position (≥5 bp from fragment ends), and local-context (≤2 nearby non-reference events) for a final set of 44 SNPs (1 homozygous, 43 heterozygous), which served as known variants for sensitivity evaluation in each dilution. Analysis of each spike-in SNP using Mutect2 demonstrated sensitivity at 1% VAF (detection of the single homozygous SNP), with 88.6% sensitivity at 0.5% and 70.5% at 0.25% (Supplementary Table 3).

For somatic mutation discovery in brain samples, SNVs were called from Hybrid-MS1, Hybrid-MS2, and Duplex consensus BAMs using Mutect2 (GATK v4.6.1.0) in single-sample mode with default parameters, including minimum base quality 30. Somatic indels were called exclusively from Duplex BAMs, and residual artefacts in repetitive regions were removed by excluding calls overlapping the hg38 RepeatMasker track. Systematic false-positive patterns identified during artificial mosaic analysis were used to inform a custom Python-based filtering pipeline (Supplementary Table 4) applied to all VCFs. Variants observed in more than one individual were excluded as potential sequencing artefacts or contamination.

All candidates were manually inspected using IGV across every region and at all three stringency levels, enabling exclusion of residual artefacts and detection of the same mutation in additional brain regions where VAFs fell below Mutect2 calling thresholds. Regardless of the stringency level at which a variant was initially called, inclusion in the final callset required evidence of a supporting duplex read in at least one brain region, confirmed by IGV inspection. To complement Mutect2-based calling, targeted read-level interrogation of 56 CH mutation coordinates compiled from CH cohort studies(27–37) (Supplementary Table 5) was performed using bcftools mpileup. Variants at the same codons supported by ≥5 duplex alternate reads in one brain region and confirmed by IGV were included in the callset, yielding two additional mutations. VAFs were calculated from IGV read counts as alternate reads / total reads, using Hybrid-MS2 BAMs for SNVs and Duplex BAMs for indels.

### Functional annotation of somatic mutations

OpenCRAVAT (Cancer-Related Analysis of Variants Toolkit, v2.15.0) was applied to annotate mutations into categories including intronic, UTR, flanking, and coding, and to provide CADD (Combined Annotation Dependent Depletion, v1.6) Phred scores for deleteriousness predictions.(38) For coding variants, functional consequences were classified based on predicted impacts on amino acid sequence. Missense variants within CH genes were additionally classified as predicted cancer drivers or passengers using CanDrA Plus (GENERAL cancer model),(39) run locally following coordinate liftover from hg38 to hg19 using pyliftover.

### Statistical Analysis

All statistical analyses were performed using Python (v3.12.2) with SciPy (v1.11) and Statsmodels (v0.14.4). As most variables deviated from normality and sample sizes were modest, non-parametric statistical tests were applied unless otherwise stated. All tests were two-tailed and statistical significance defined as p < 0.05, with nominal values provided.

Continuous variables (post-mortem interval, and sequencing depth) were compared between MSA and controls using Mann-Whitney U tests.

To compare total mutations and CH mutations between groups whilst accounting for age, ordinary least squares (OLS) linear regression models were constructed with mutation number as the dependent variable and disease group and age at death as independent variables. Separate models were constructed for combined MSA cases and for each MSA subtype.

### Data Sharing

The data that support the findings of this study are available on request from the corresponding author, subject to formal approval by QSBB. All code used for somatic variant calling and filtering is available at https://github.com/b-thompson99/XT-HS2-Small-Somatic.

## RESULTS

### Low-frequency somatic mutations are detectable in MSA and control brains

We performed duplex sequencing of target genes (*SNCA*, *KCTD7*, and ten cancer driver genes), applying UMI-based error correction to enhance variant detection (Fig. 1B). Our sequencing approach generated consensus reads at three stringency levels: Duplex, Hybrid-MS2, and Hybrid-MS1, with respective mean depths of 963×, 2,171×, and 7,249× (Supplementary Table 6). MSA and control groups showed no significant differences in sequencing depth across all stringency levels and brain regions (all p>0.05, Mann-Whitney U tests). Following alignment, variant calling, filtering, and targeted interrogation of known CH mutation sites, all candidate variants underwent comprehensive IGV inspection (Fig. 1C; see methods). Analysis was conducted only on high-confidence variants supported by duplex consensus reads. The final callset consisted of 34 variants comprising 17 SNVs, 16 deletions, and one insertion (Supplementary Table 7).

### MSA brains harbor elevation of somatic clonal hematopoiesis gene mutations

The callset was dominated by somatic mutations in cancer driver genes (Fig. 2A). Focusing on the seven CH-associated genes, we detected 14 mutations across all brains, 12 in MSA and two in controls. Of these, 10 led to coding changes (Table 1; IGV traces in Supplementary Fig. 4), eight of which were in MSA. Each MSA brain harbored on average 0.6 CH mutations, against 0.22 in controls. Employing age-adjusted linear regression, CH mutation number at the whole brain level appeared higher in MSA, though this did not reach significance (median [IQR]: 1 [0–1] vs 0 [0–0]; p=0.206; (Supplementary Fig. 3). We detected at least one CH mutation in 10/20 MSA brains (six SND and four OPCA), versus 2/9 controls. As we had three brain regions in all brains (except one where we had two), we compared the prevalence of multi-regional CH mutation, which were present in six MSA patients, but no controls. Indeed, three MSA patients harbored CH mutations in all three brain regions.

**Figure 2.**
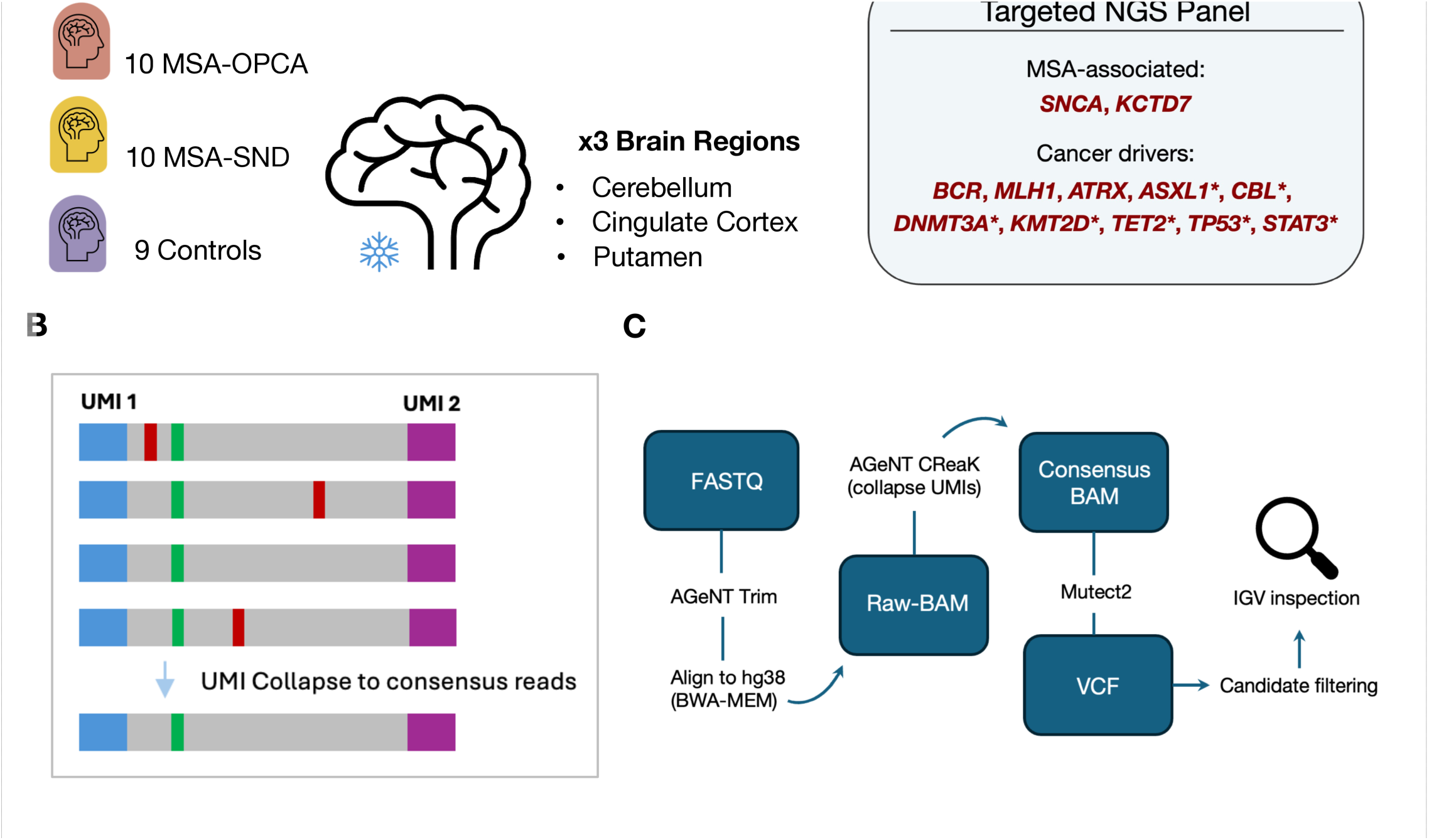
Somatic CH mutations in MSA and control brains. **(A) Oncoprint showing CH gene mutations across all individuals:** MSA subtypes (OPCA, SND) and controls indicated. Each column represents one individual; half-cell denotes individual with two variants in one gene **(B) Age-adjusted OLS regression of CH mutation**s per individual across cerebellum, cingulate cortex, and putamen separately. Comparisons shown between MSA-OPCA, MSA-SND, and controls; p-values indicated per panel.

**Table 1.** Non-synonymous clonal hematopoiesis variants detected in MSA patients and controls.

| Variant ID | Mutation | Case ID | Consequence | Protein Change | CADD-Phred | CanDrA | Variant allele fraction (%) |  |  |
| --- | --- | --- | --- | --- | --- | --- | --- | --- | --- |
|  |  |  |  |  |  |  | CER | CC | PUT |
| *DNMT3A*_SNV2 | chr2:g.25240708T>C | SND-04 | Missense | p.Asp702Gly | 30.0 | Driver (3.511) | 0.93 | 2.17 | 0.88 |
| *DNMT3A*_SNV3 | chr2:g.25241582C>T | SND-09 | Missense | p.Arg688Cys | 31.0 | Driver (3.475) | - | 0.21 | 0.15 |
| *DNMT3A*_DEL1 | chr2:g.25247067_25247081 del | SND-08 | In-frame | p.Met364_Ala368 del | 22.5 | - | 0.38 | 0.35 | 0.54 |
| *KMT2D*_SNV3 | chr12:g.49034200C>T | SND-01 | Missense | p.Arg3536His | 27.6 | Driver (3.644) | - | 0.40 | 0.34 |
| *KMT2D*_DEL1 | chr12:g.49024674_49024677 del | SND-15 | Frameshift | p.Leu5318Ser fs Ter14 | 36.0 | - | - | 0.40 | - |
| *KMT2D*_DEL2 | chr12:g.49033837_49033839 del | OPCA-05 | In-frame | p.Gln3623del | 18.2 | - | - | - | 0.67 |
| *TET2*_SNV1 | chr4:g.105275662G>T | OPCA-01 | Missense | p.Val1718Leu | 1.1 | Driver (0.796) | 0.02 | 0.29 | 0.05 |
| *TP53*_SNV2 | chr17:g.7674221G>A | SND-06 | Missense | p.Arg248Trp | 29.2 | Driver (30.671) | 0.39 | 0.39 | - |
| *CBL*_SNV2 | chr11:g.119278528T>C | Control -05 | Missense | p.Cys416Arg | 29.9 | Passenger (-0.618) | 1.47 | - | - |
| *DNMT3A*_SNV1 | chr2:g.25240652T>A | Control -08 | Stop gained | p.Lys721Ter | 42.0 | - | 1.02 | - | - |

We next asked whether CH mutation number varied across individual brain regions, comparing each between MSA and controls (Fig. 2B). MSA cases had more in the cortex compared to controls (p=0.033), driven predominantly by SND (p=0.011). SND brains also demonstrated significant elevation of CH mutations in the putamen, their most affected region, versus controls (p=0.034). OPCA cases showed a consistent though non-significant trend towards CH elevation across all three brain regions.

### Somatic brain mutations in CH genes include protein-altering events with predicted oncogenic potential

To contextualize the functional significance of CH mutations detected, we cross-referenced our callset against a compiled panel of 56 CH mutation positions derived from CH cohort studies (see Methods). As no dedicated CH-specific variant annotation tool currently exists, we also applied CanDrA Plus (GENERAL pan-cancer model) to all missense CH variants, classifying mutations as predicted cancer driver or passenger based on integrated structural, evolutionary and gene-level features. CADD scores were obtained to provide complementary evidence of deleteriousness. Of the 10 coding CH variants (Table 1), three MSA variants occurred at residues reported as recurrently mutated in CH. Two of these were *DNMT3A* missense variants in SND cases: DNMT3A_SNV2 in SND-04, located between the catalytic loop and substrate binding motif of the methyltransferase domain, detected in all three brain regions and exhibiting the highest VAF of any CH variant in the callset; and DNMT3A_SNV3 in SND-09, another observed CH substitution within the same domain, detected in cortex and putamen. Both were classified as predicted drivers by CanDrA. The third MSA variant at a recurrent CH residue was TP53_SNV2 in SND-06, detected in cerebellum and cortex, and is an established CH hotspot; TP53_SNV2 received the highest CanDrA driver score of any variant in the callset.

Beyond those found at previously confirmed CH coordinates, a further five coding CH variants were identified in MSA brains; the two missense variants among these were both predicted drivers by CanDrA. TET2_SNV1 in OPCA-01 was detected in all three brain regions; and KMT2D_SNV3 in SND-01 was present in the cortex and putamen. Three additional coding variants were indels: in *DNMT3A*, DNMT3A_DEL1 (p.Met364_Ala368del) removed five amino acids from the PWWP domain, a chromatin-targeting domain essential for *DNMT3A* localization to heterochromatin and DNA methylation activity, detected across all three brain regions of SND-08; in *KMT2D*, KMT2D_DEL1, a frameshift deletion predicted to ablate the C-terminal SET domain and result in loss of methyltransferase activity through nonsense-mediated decay, detected exclusively in the cortex of SND-15; and KMT2D_DEL2 (p.Gln3623del), an in-frame deletion restricted to the putamen of OPCA-05. The remaining two coding CH mutations were both detected in a single brain region of control brains: CBL_SNV2 in Control-05, and DNMT3A_SNV1 in Control-08. Both were detected in individuals aged 88, older than all MSA patients and all but one control, potentially reflecting the age-dependence of CH acquisition. Both were restricted to the cerebellum with VAF >1%, whereas six of eight coding mutations in MSA cases showed presence across two or more regions, supporting a more widespread disease-associated process regarding functional CH clones in the MSA brain.

### Small somatic mutations in *SNCA* and *KCTD7* do not appear to play a major role in MSA

Despite comprehensive coverage of the *SNCA* exons, no coding mutations were detected in any MSA patient or control, though seven non-coding *SNCA* mutations were identified (two in MSA, five in controls). The only one found in all brain regions was SNCA_SNV5 (chr4:g.89838202T>C), in SND-01, with the highest VAF (0.77%) in the putamen. This variant was in *SNCA-AS1*, within a region associated with LBD.(25) In *KCTD7*, we detected a total of four unique mutations, two deletions and two SNVs. Both deletions were in MSA brains, including one coding in-frame deletion (KCTD7_DEL2, p.Val4del) predicted deleterious (CADD-Phred=20.7), which was, however, absent in the cerebellum of the OPCA case carrying it. Both SNVs, including one missense predicted deleterious (CADD-Phred=19.8), were detected in control brains. We therefore find no evidence that *SNCA* or *KCTD7* somatic SNVs and indels broadly underlie MSA.

## DISCUSSION

In this work, we performed deep targeted sequencing to detect small somatic mutations (SNVs and indels) in a selected gene set in MSA brains. We used UMI-based error correction with duplex sequencing and curation based on analysis of artificial mosaics to minimize the risk of false positives, with 88.6% sensitivity at 0.5% VAF. We hypothesized that somatic indels within *SNCA* may contribute to MSA, as we have already demonstrated a clear relationship of somatic *SNCA* CNVs (gains) to MSA and PD.(8–11) In previous work to detect *SNCA* somatic SNVs in synucleinopathies, we did not detect any, but only three MSA cases were included.(16) In the current study, despite including key brain regions affected differentially in SND and OPCA, we found no small somatic coding mutations within *SNCA*, and we cannot draw any conclusion from the multi-region mutation in *SNCA-AS1* in one case. We also included *KCTD7*, a novel genetic risk for MSA.(5) This too revealed no excess somatic mutations in MSA. Small somatic mutations in these genes therefore appear unlikely to underlie MSA, although rare events, intronic mutations, initiating mutations in cells which have subsequently died, and the impact of variants below our detection thresholds, cannot be excluded.

Focusing on genes involved in CH, we identified excess somatic mutations in MSA brains. Overall, all regions in MSA had either significantly larger CH mutation number compared to the same region in controls, or a similar trend. The fact that six patients (30%) but no controls harbored CH mutations in more than one brain region, and in all three regions for three of them, also implies a widespread presence of CH clones throughout the MSA brain. In contrast, the two control coding CH mutations were only in the cerebellum, despite a VAF >1%, suggesting a more localized process. Furthermore, both were in individuals six years older than the oldest MSA cases. Further analysis of somatic coding CH mutations revealed five MSA mutations predicted to be cancer drivers, three of which occurred at residues recurrently mutated in prior CH cohorts, suggestive of preferential expansion of the clone carrying them; the single missense CH mutation in controls was predicted to be a passenger.

Several independent findings support the relevance of our results. CH with *DNMT3A* mutations was elevated in the blood of 100 MSA patients compared to controls.(21) Prior work examining somatic mutations in brains from patients with AD and LBD detected non-synonymous CH mutations in 40% of LBD cases versus 7% of controls, suggesting CH presence in the brain may be a broader feature of synucleinopathies.(40) This may also have been an underestimate, since only 27 CH genes were studied. More recently, somatic mutations in CH driver genes were identified in AD brains, where they demonstrated evidence of positive selection.(41) Mutations in six CH genes were confirmed to be in microglia, with mutant cells constituting 20-40% of microglia. In addition to relevant gene mutations, mosaic chromosomal alterations (mCA) are a feature of CH, and were also found in Alzheimer’s microglia.(42) Furthermore, another AD study correlated blood mCA with matched brain single nucleus RNAseq, revealing microglia-annotated cells in brain with the same mCA.(42) Notably, both studies reported aberrant transcriptional profiles of microglia with mCA.

Overall, if the detected CH mutations are indeed in microglia as expected, mutant microglia may drive pathology through inflammatory mechanisms. Hematopoietic expression of a common human CH mutation rendered monocyte-derived microglia pathogenic in an aging mouse model, where they accumulated in selected brain regions and promoted motor deficits resembling atypical Parkinsonian disorders.(43) An older MSA mouse model showed early progressive microglial activation in substantia nigra which correlated with dopaminergic neuronal loss, and microglial suppression was protective.(44) In the human brain, proliferating mutant microglia were reported in rare histiocytosis-associated neurodegeneration.(45) These findings suggest CH-mutant monocytes can infiltrate the brain and adopt a microglial phenotype as discussed.(41) A hyperreactive pro-inflammatory phenotype could potentiate MSA-associated neuroinflammation and degeneration. Neuroinflammation is a consistent feature of MSA, although whether it acts as a primary driver of degeneration, a secondary response to proteinopathy, or perhaps both, remains uncertain.(46,47) Accumulating evidence suggests misfolded α-synuclein triggers microglial activation and astrogliosis, in turn accelerating α-synuclein aggregation and demyelination. The resulting inflammatory response to oligodendroglial α-synuclein pathology, marked by microglial activation and chemokine upregulation,(48) may establish a more permissive environment for peripheral immune cell entry. Indeed, peripherally derived immune cells were significantly elevated in post-mortem MSA brain tissue, adopted microglial function, and appeared to accumulate surrounding GCIs in the putamen and directly on top of dopaminergic neurons in the substantia nigra.(49) In a mouse model overexpressing α-synuclein in oligodendrocytes, a reduction in neuroinflammation and demyelination was observed following knockout of T cells.(49) Together, these findings support a pathogenic role for infiltrating peripheral immune cells in the MSA brain. The converse, however, can’t be excluded, with *TET2*-mutant myeloid cells protective in an AD mouse model.(50) Indeed, whether CH clones represent active contributors to neurodegeneration, secondary consequences of disease processes, or bystanders in MSA pathogenesis remains to be determined.

The limitations of our work are that we only studied a subset of CH genes, albeit enriched for the most mutated, the lack of direct demonstration of the cell type(s) harboring mutations, and the absence of matched peripheral blood. Whilst blood contamination could in theory be the source of brain CH mutations, several findings argue against this. If blood contamination were responsible, VAF would be expected to correlate with vascular density, while the patterns across regions are very variable (Table 1). In previous LBD and AD work, CH mutation VAF was consistently higher in matched peripheral blood, where available, than brain, but the brain VAF was higher than expected from blood contamination alone.(40) Whilst our sequencing approach could not distinguish cell types, the hypothesis that an elevation of CH in the MSA brain reflects mutations within monocyte derived cells indistinguishable from CNS-resident microglia, rather than neurons or oligodendrocytes, is supported by multiple lines of evidence. Firstly, the enrichment of *DNMT3A* mutations in peripheral blood from MSA patients suggests CH elevation may originate from bone marrow-derived hematopoiesis. Beyond this, the elevation of CH mutations in AD brains is likely also derived from cells of bone marrow hematopoietic origin considering the 100% detection rate and greater VAF of mutations in matched blood.

In summary, we provide evidence that CH mutations are elevated in MSA brains, implicating a potential role for somatic mutational processes beyond *SNCA* CNVs. Multi-regional detection patterns are consistent with widespread clonal proliferation or infiltration of mutation-harboring cells, which have presumed hematopoietic origin. Future sequencing of DNA derived from individual cell types will confirm the cell type(s) harboring these mutations, and functional studies will be required to elucidate possible mechanistic contributions of CH to MSA pathogenesis. With future translation in mind, a peripheral origin of CH in MSA may offer therapeutic opportunities distinct from targeting CNS-resident pathology, including strategies to prevent pathogenic monocyte infiltration, or their modulation, which has shown promise in animal studies.(44,49) Moreover, peripheral CH burden may serve as an accessible biomarker in MSA.

## Data Availability

The data that support the findings of this study are available on request from the corresponding author, subject to formal approval by the Queen Square Brain Bank. The data are not publicly available due to ethics and consent restrictions.

## ACKNOWLEDGEMENTS

This work was funded by Mission MSA (formerly the MSA Coalition) and the MSA Trust. We are grateful to Queen Square Brain Bank and the individuals who donated their brains. The authors acknowledge support from the Biotechnology and Biological Sciences Research Council (BBSRC), part of UK Research and Innovation, Core Capability Grant BB/CCG2220/1 at the Earlham Institute. Part of this work was delivered via Transformative Genomics, the BBSRC funded National Bioscience Research Infrastructure (BBS/E/ER/23NB0006) at Earlham Institute, by members of the Technical Genomics Pipelines and Core Bioinformatics Groups. Artificial intelligence tools (Claude, Anthropic) were used to assist in the development of custom Python scripts for bioinformatic filtering and statistical analysis. All code was reviewed and validated by the authors.

## AUTHORS’ ROLES

Design: CP, ZJ, EG. Supervision: CP. Experimental work: DH, CM. Analysis: BT. Drafting manuscript: BT. Manuscript editing: all authors.

## FINANCIAL DISCLOSURES

CP received grants from Mission MSA (formerly the MSA Coalition) and the MSA Trust for the study of somatic mutations in MSA. The other authors have no disclosures.

## Notes

### Competing Interest Statement

The authors have declared no competing interest.

### Funding Statement

This work was funded by the Mission MSA (formerly the MSA Coalition) and MSA Trust.

### Author Declarations

All donors or their next of kin had given informed consent for the use of their brain in this research, and the study was approved by NHS Health Research Authority Ethics Committee London-Central (REC reference 23/LO/0044).

### Summary of Updates

The main change is the increased stringency of the mutation calls. Only calls supported by duplex sequencing are included, which results in a smaller callset (72% of original calls), which is more robust, with appropriate statistical adjustments.

