## Supplementary Figure for "Multiple system atrophy is associated with brain somatic mutations in clonal hematopoiesis genes"

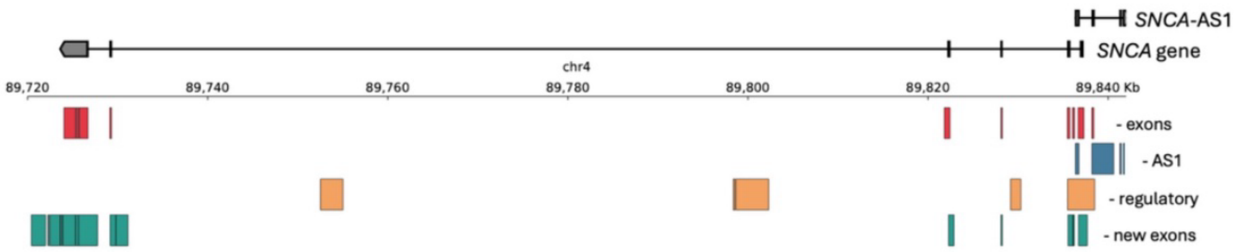


**Supplementary Figure 1. *SNCA* locus target map showing sequencing coverage**
Genomic structure of the *SNCA* locus on chromosome 4 showing all targeted regions including coding exons, 10 bp flanking sequence for all transcribed regions, enhancer and promoter regions, 15 low-abundance alternative exons, and *SNCA-AS1*. Full target coordinates are provided in Supplementary Table 2.

| *DNMT3A*_SNV2 | chr2:g.25240708T>C |
|---|---|

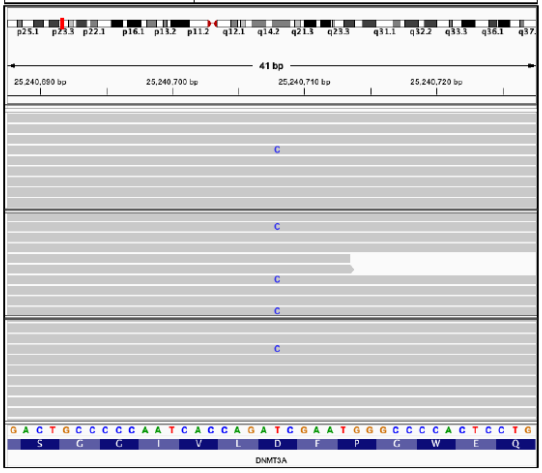


| *DNMT3A*_SNV3 | chr2:g.25241582C>T |
|---|---|

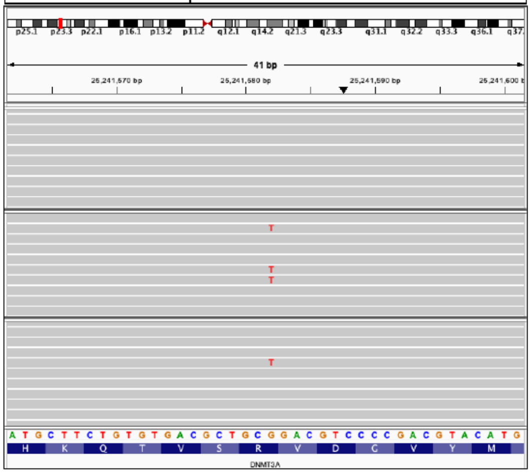


| *DNMT3A*_DEL1 | chr2:g.25247067_25247081del |
|---|---|

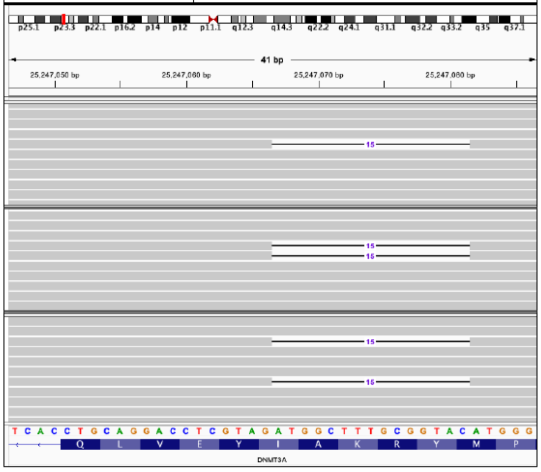


| *KMT2D*_SNV3 | chr12:g.49034200C>T |
|---|---|

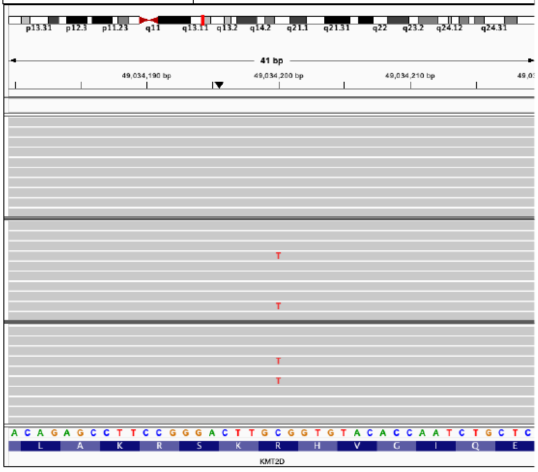


| *KMT2D*_DEL1 | chr12:g.49024674_49024677del |
|---|---|

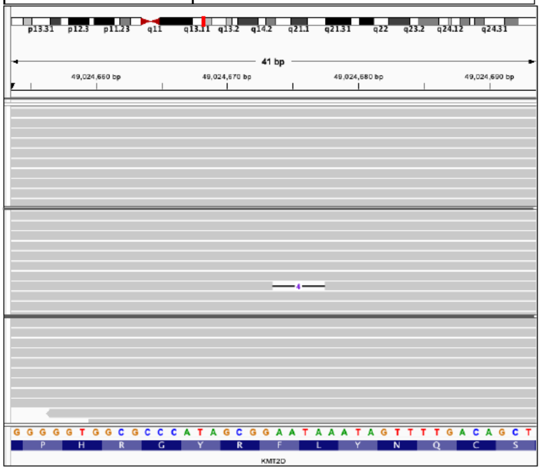


| *KMT2D*_DEL2 | chr12:g.49033837_49033839del |
|---|---|

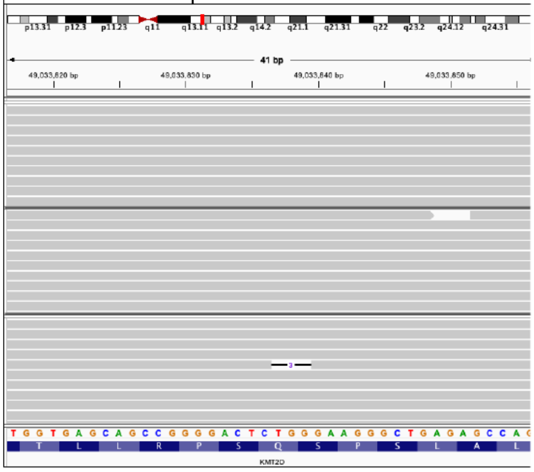

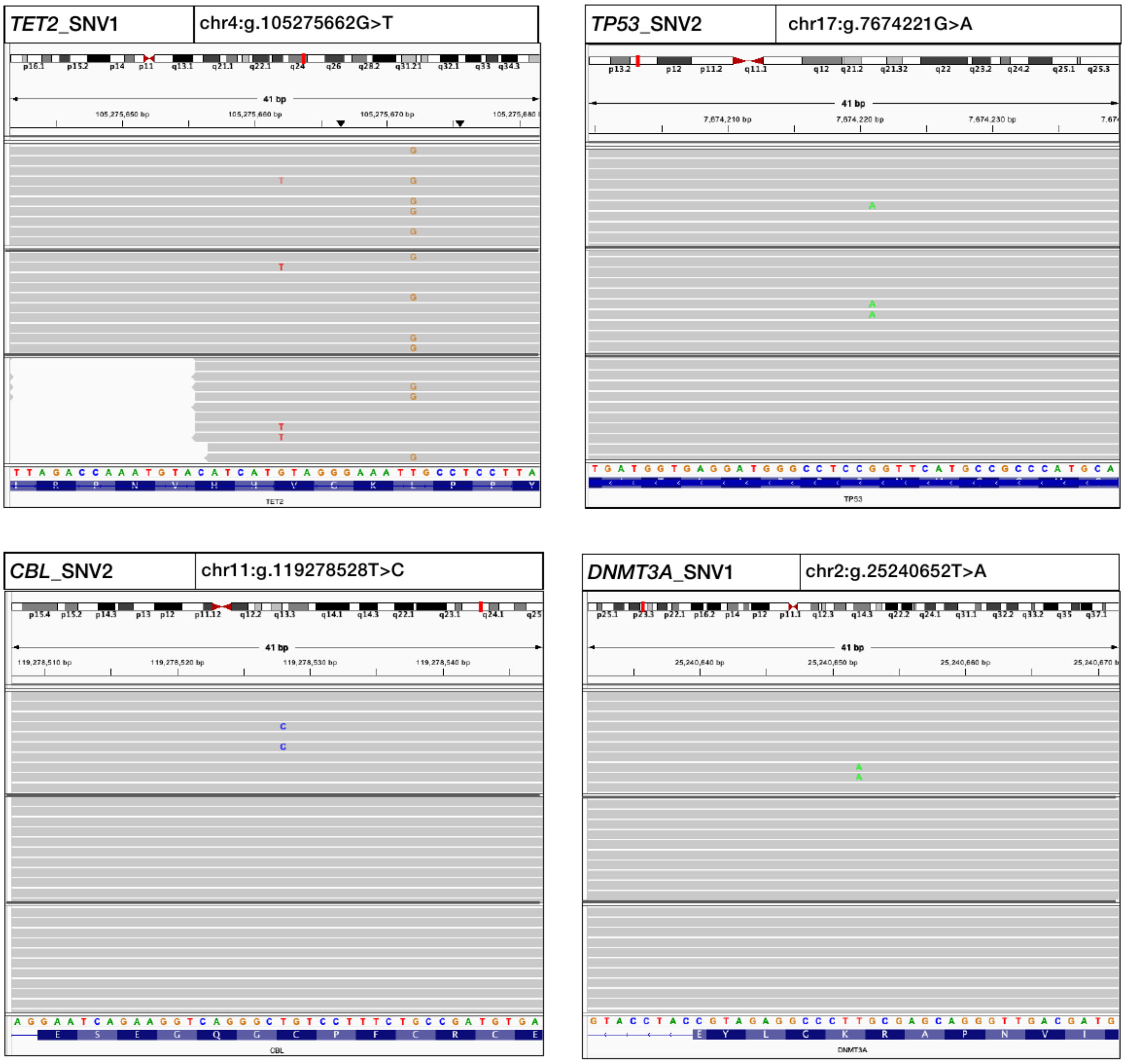


**Supplementary Figure 2. IGV traces of coding CH mutations detected in MSA and control brains**

Each panel shows the variant ID and chromosomal position in the header. Colored bases indicate the non-reference alternate allele; gray reads indicate the reference allele. For SNVs, Hybrid-MS2 consensus reads are displayed; for indels, full duplex consensus reads are displayed. Three brain regions are shown per variant: cerebellum (top), cortex (middle), and putamen (bottom). Variant details in Table 1.

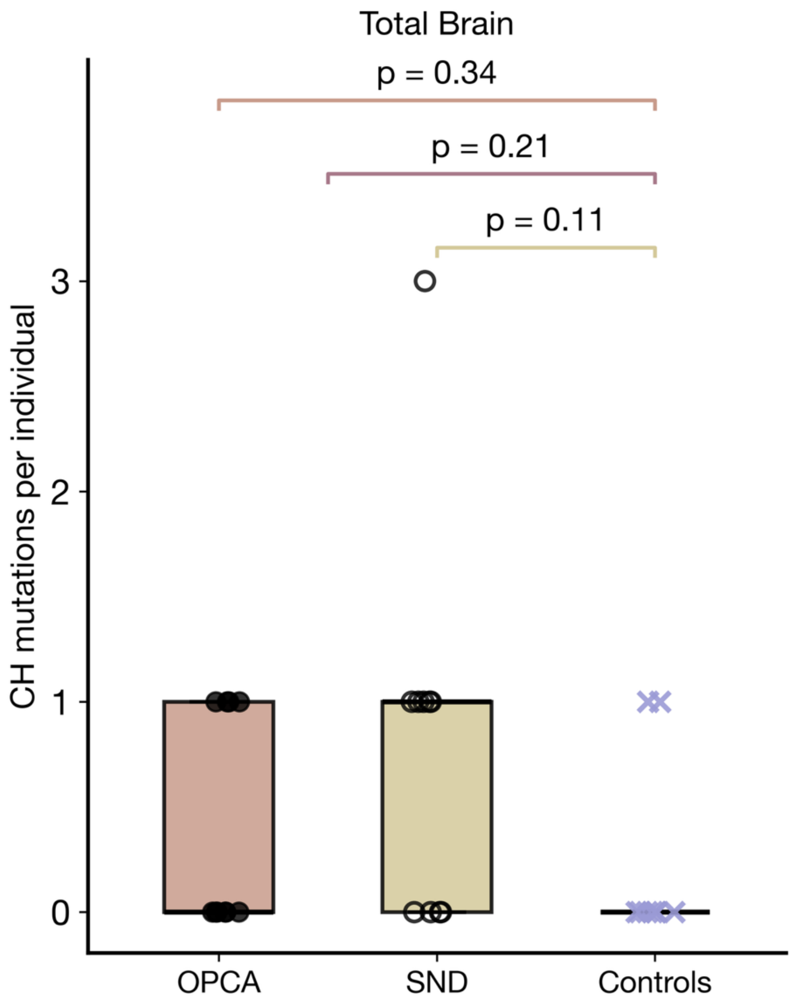


**Supplementary Figure 3. Whole brain CH mutations across MSA subtypes and controls**

Total CH mutations per individual across all three brain regions combined in MSA-OPCA, MSA-SND, and controls. Comparisons performed using age-adjusted ordinary least squares regression. Box plots show median (center line), interquartile range (box), and range (whiskers); individual data points are overlaid. Bracket colors indicate comparisons: orange, MSA-OPCA versus controls; red, combined MSA versus controls; yellow, MSA-SND versus controls.
