## Supplementary Table for "Multiple system atrophy is associated with brain somatic mutations in clonal hematopoiesis genes"

#### Supplementary Table 1. Patient demographics

| Individual ID | Age at death (completed decades) | PMI (hours) |
|---|---|---|
| OPCA-01 | 6 | 81.2 |
| OPCA-02 | 7 | 64.1 |
| OPCA-03 | 6 | 46.25 |
| OPCA-05 | 5 | 88.1 |
| OPCA-06 | 6 | 62.7 |
| OPCA-07 | 6 | 60.5 |
| OPCA-08 | 7 | 28.7 |
| OPCA-09 | 7 | 85.3 |
| OPCA-11 | 5 | 88.4 |
| OPCA-12 | 5 | 52 |
| SND-01 | 7 | 34 |
| SND-04 | 6 | 62.25 |
| SND-05 | 8 | 41 |
| SND-06 | 5 | 45 |
| SND-07 | 7 | 107 |
| SND-08 | 6 | 28.7 |
| SND-09 | 8 | 43.75 |
| SND-13 | 6 | 79 |
| SND-15 | 6 | 73.5 |
| SND-16 | 6 | 44.75 |
| Control-02 | 7 | 47 |
| Control-03 | 5 | — |
| Control-04 | 7 | 101.3 |
| Control-05 | 8 | 11.1 |
| Control-06 | 9 | 46.3 |
| Control-07 | 6 | 44.55 |
| Control-08 | 8 | 16.15 |
| Control-11 | 7 | 38.5 |
| Control-14 | 7 | 76.1 |

OPCA = olivopontocerebellar atrophy; SND = striatonigral degeneration; PMI = post-mortem interval. All patients had cerebellum, cingulate cortex, and putamen sampled except OPCA-02 (no cortex available). Age at death is given per completed decade, ie 6 indicates age 60-69. No significant difference was detected for PMI ($p = 0.252$).

**Supplementary Table 2. Target panel coordinates**

| Gene | Chromosome | Start | End | Size (bp) |
|---|---|---|---|---|
| *SNCA (new exon)* | chr4 | 89720461 | 89727756 | 7,295 |
| *SNCA* | chr4 | 89724048 | 89726710 | 2,662 |
| *SNCA* | chr4 | 89729143 | 89729327 | 184 |
| *SNCA (new exon)* | chr4 | 89729194 | 89731158 | 1,964 |
| *SNCA (new exon)* | chr4 | 89752547 | 89754996 | 2,449 |
| *SNCA (new exon)* | chr4 | 89798377 | 89802296 | 3,919 |
| *SNCA* | chr4 | 89821798 | 89822438 | 640 |
| *SNCA (new exon)* | chr4 | 89822246 | 89822981 | 735 |
| *SNCA* | chr4 | 89828092 | 89828234 | 142 |
| *SNCA (new exon)* | chr4 | 89828143 | 89828194 | 51 |
| *SNCA (new exon)* | chr4 | 89829189 | 89830284 | 1,095 |
| *SNCA* | chr4 | 89835496 | 89835745 | 249 |
| *SNCA (new exon)* | chr4 | 89835512 | 89838475 | 2,963 |
| *SNCA (new exon)* | chr4 | 89835547 | 89836088 | 541 |
| *SNCA* | chr4 | 89836076 | 89836263 | 187 |
| *SNCA (new exon)* | chr4 | 89836127 | 89836242 | 115 |
| *SNCA* | chr4 | 89836692 | 89837278 | 586 |
| *SNCA (new exon)* | chr4 | 89836743 | 89837672 | 929 |
| *SNCA* | chr4 | 89838201 | 89838365 | 164 |
| *SNCA-AS1* | chr4 | 89836390 | 89836744 | 354 |
| *SNCA-AS1* | chr4 | 89838250 | 89840605 | 2,355 |
| *SNCA-AS1* | chr4 | 89841330 | 89841410 | 80 |
| *SNCA-AS1* | chr4 | 89841700 | 89842001 | 301 |
| *KCTD7* | chr7 | 66628756 | 66629218 | 462 |
| *KCTD7* | chr7 | 66633264 | 66633454 | 190 |
| *KCTD7* | chr7 | 66638242 | 66644025 | 5,783 |
| *KCTD7* | chr7 | 66647867 | 66649077 | 1,210 |
| *ASXL1* | chr20 | 32358319 | 32358842 | 523 |
| *ASXL1* | chr20 | 32358955 | 32359861 | 906 |
| *ASXL1* | chr20 | 32360006 | 32360596 | 590 |
| *ASXL1* | chr20 | 32360623 | 32360816 | 193 |
| *ASXL1* | chr20 | 32366373 | 32366477 | 104 |
| *ASXL1* | chr20 | 32367716 | 32367739 | 23 |
| *ASXL1* | chr20 | 32369001 | 32369133 | 132 |
| *ASXL1* | chr20 | 32371767 | 32371884 | 117 |

| Gene | Chromosome | Start | End | Size (bp) |
|---|---|---|---|---|
| *ASXL1* | chr20 | 32372153 | 32372563 | 410 |
| *ASXL1* | chr20 | 32427061 | 32428432 | 1,371 |
| *ASXL1* | chr20 | 32428753 | 32431494 | 2,741 |
| *ASXL1* | chr20 | 32431545 | 32431689 | 144 |
| *ASXL1* | chr20 | 32432348 | 32432995 | 647 |
| *ASXL1* | chr20 | 32433273 | 32439329 | 6,056 |
| *ATRX* | chrX | 77504867 | 77508639 | 3,772 |
| *ATRX* | chrX | 77520777 | 77522478 | 1,701 |
| *ATRX* | chrX | 77523241 | 77523411 | 170 |
| *ATRX* | chrX | 77557440 | 77557655 | 215 |
| *ATRX* | chrX | 77558658 | 77558856 | 198 |
| *ATRX* | chrX | 77561636 | 77561718 | 82 |
| *ATRX* | chrX | 77574239 | 77574368 | 129 |
| *ATRX* | chrX | 77575543 | 77575615 | 72 |
| *ATRX* | chrX | 77575682 | 77575763 | 81 |
| *ATRX* | chrX | 77589823 | 77589950 | 127 |
| *ATRX* | chrX | 77593685 | 77596885 | 3,200 |
| *ATRX* | chrX | 77599400 | 77599590 | 190 |
| *ATRX* | chrX | 77599721 | 77599830 | 109 |
| *ATRX* | chrX | 77600423 | 77600765 | 342 |
| *ATRX* | chrX | 77616234 | 77616521 | 287 |
| *ATRX* | chrX | 77616602 | 77616740 | 138 |
| *ATRX* | chrX | 77618795 | 77618991 | 196 |
| *ATRX* | chrX | 77620384 | 77620542 | 158 |
| *ATRX* | chrX | 77633196 | 77633394 | 198 |
| *ATRX* | chrX | 77633555 | 77633875 | 320 |
| *ATRX* | chrX | 77634411 | 77634713 | 302 |
| *ATRX* | chrX | 77635904 | 77636066 | 162 |
| *ATRX* | chrX | 77651972 | 77652363 | 391 |
| *ATRX* | chrX | 77654087 | 77654210 | 123 |
| *ATRX* | chrX | 77656549 | 77656663 | 114 |
| *ATRX* | chrX | 77663371 | 77663568 | 197 |
| *ATRX* | chrX | 77664634 | 77664788 | 154 |
| *ATRX* | chrX | 77673943 | 77676308 | 2,365 |
| *ATRX* | chrX | 77681509 | 77684603 | 3,094 |
| *ATRX* | chrX | 77684928 | 77685016 | 88 |
| *ATRX* | chrX | 77688807 | 77688937 | 130 |

| Gene | Chromosome | Start | End | Size (bp) |
|---|---|---|---|---|
| *ATRX* | chrX | 77690792 | 77690935 | 143 |
| *ATRX* | chrX | 77691214 | 77691301 | 87 |
| *ATRX* | chrX | 77693813 | 77693947 | 134 |
| *ATRX* | chrX | 77696566 | 77696714 | 148 |
| *ATRX* | chrX | 77697572 | 77697645 | 73 |
| *ATRX* | chrX | 77698563 | 77698639 | 76 |
| *ATRX* | chrX | 77699499 | 77699557 | 58 |
| *ATRX* | chrX | 77717120 | 77717253 | 133 |
| *ATRX* | chrX | 77785834 | 77786279 | 445 |
| *BCR* | chr22 | 23179693 | 23179842 | 149 |
| *BCR* | chr22 | 23180199 | 23182249 | 2,050 |
| *BCR* | chr22 | 23198242 | 23198395 | 153 |
| *BCR* | chr22 | 23199200 | 23199362 | 162 |
| *BCR* | chr22 | 23242807 | 23242995 | 188 |
| *BCR* | chr22 | 23251021 | 23251250 | 229 |
| *BCR* | chr22 | 23253788 | 23253990 | 202 |
| *BCR* | chr22 | 23260939 | 23261064 | 125 |
| *BCR* | chr22 | 23261344 | 23261550 | 206 |
| *BCR* | chr22 | 23268397 | 23268525 | 128 |
| *BCR* | chr22 | 23271521 | 23271602 | 81 |
| *BCR* | chr22 | 23273070 | 23273143 | 73 |
| *BCR* | chr22 | 23273623 | 23273784 | 161 |
| *BCR* | chr22 | 23283198 | 23284108 | 910 |
| *BCR* | chr22 | 23285022 | 23285211 | 189 |
| *BCR* | chr22 | 23287148 | 23287288 | 140 |
| *BCR* | chr22 | 23288086 | 23288182 | 96 |
| *BCR* | chr22 | 23289506 | 23291141 | 1,635 |
| *BCR* | chr22 | 23292530 | 23292648 | 118 |
| *BCR* | chr22 | 23295013 | 23295165 | 152 |
| *BCR* | chr22 | 23302330 | 23302873 | 543 |
| *BCR* | chr22 | 23306143 | 23306259 | 116 |
| *BCR* | chr22 | 23306734 | 23306840 | 106 |
| *BCR* | chr22 | 23307557 | 23307774 | 217 |
| *BCR* | chr22 | 23309413 | 23310443 | 1,030 |
| *BCR* | chr22 | 23311686 | 23311928 | 242 |
| *BCR* | chr22 | 23312377 | 23313031 | 654 |
| *BCR* | chr22 | 23313957 | 23314083 | 126 |

| Gene | Chromosome | Start | End | Size (bp) |
|---|---|---|---|---|
| *BCR* | chr22 | 23314541 | 23314724 | 183 |
| *BCR* | chr22 | 23315422 | 23318047 | 2,625 |
| *CBL* | chr11 | 119206265 | 119206622 | 357 |
| *CBL* | chr11 | 119232437 | 119232705 | 268 |
| *CBL* | chr11 | 119271724 | 119271891 | 167 |
| *CBL* | chr11 | 119273857 | 119274034 | 177 |
| *CBL* | chr11 | 119274821 | 119274963 | 142 |
| *CBL* | chr11 | 119275986 | 119276144 | 158 |
| *CBL* | chr11 | 119277746 | 119277854 | 108 |
| *CBL* | chr11 | 119278155 | 119278307 | 152 |
| *CBL* | chr11 | 119278499 | 119278723 | 224 |
| *CBL* | chr11 | 119284958 | 119285110 | 152 |
| *CBL* | chr11 | 119285178 | 119285576 | 398 |
| *CBL* | chr11 | 119287841 | 119287956 | 115 |
| *CBL* | chr11 | 119296907 | 119297044 | 137 |
| *CBL* | chr11 | 119297373 | 119297491 | 118 |
| *CBL* | chr11 | 119298347 | 119298550 | 203 |
| *CBL* | chr11 | 119299484 | 119308159 | 8,675 |
| *CBL* | chr11 | 119313407 | 119313936 | 529 |
| *DNMT3A* | chr2 | 25227844 | 25234430 | 6,586 |
| *DNMT3A* | chr2 | 25235695 | 25235835 | 140 |
| *DNMT3A* | chr2 | 25236919 | 25237015 | 96 |
| *DNMT3A* | chr2 | 25239119 | 25239225 | 106 |
| *DNMT3A* | chr2 | 25239304 | 25239523 | 219 |
| *DNMT3A* | chr2 | 25240291 | 25240460 | 169 |
| *DNMT3A* | chr2 | 25240629 | 25240740 | 111 |
| *DNMT3A* | chr2 | 25241551 | 25242025 | 474 |
| *DNMT3A* | chr2 | 25243887 | 25243992 | 105 |
| *DNMT3A* | chr2 | 25244144 | 25244348 | 204 |
| *DNMT3A* | chr2 | 25244529 | 25244662 | 133 |
| *DNMT3A* | chr2 | 25245242 | 25245342 | 100 |
| *DNMT3A* | chr2 | 25246009 | 25246074 | 65 |
| *DNMT3A* | chr2 | 25246149 | 25246319 | 170 |
| *DNMT3A* | chr2 | 25246609 | 25246786 | 177 |
| *DNMT3A* | chr2 | 25247039 | 25247576 | 537 |
| *DNMT3A* | chr2 | 25247580 | 25247909 | 329 |
| *DNMT3A* | chr2 | 25248026 | 25248262 | 236 |

| Gene | Chromosome | Start | End | Size (bp) |
|---|---|---|---|---|
| *DNMT3A* | chr2 | 25249607 | 25249734 | 127 |
| *DNMT3A* | chr2 | 25251901 | 25252141 | 240 |
| *DNMT3A* | chr2 | 25252183 | 25252325 | 142 |
| *DNMT3A* | chr2 | 25274930 | 25275097 | 167 |
| *DNMT3A* | chr2 | 25275489 | 25275553 | 64 |
| *DNMT3A* | chr2 | 25281441 | 25282721 | 1,280 |
| *DNMT3A* | chr2 | 25300128 | 25300253 | 125 |
| *DNMT3A* | chr2 | 25313902 | 25314171 | 269 |
| *DNMT3A* | chr2 | 25341815 | 25341935 | 120 |
| *DNMT3A* | chr2 | 25342419 | 25342600 | 181 |
| *KMT2D* | chr12 | 49018964 | 49021882 | 2,918 |
| *KMT2D* | chr12 | 49022032 | 49022161 | 129 |
| *KMT2D* | chr12 | 49022269 | 49022363 | 94 |
| *KMT2D* | chr12 | 49022579 | 49022885 | 306 |
| *KMT2D* | chr12 | 49024042 | 49024110 | 68 |
| *KMT2D* | chr12 | 49024567 | 49024718 | 151 |
| *KMT2D* | chr12 | 49024799 | 49024956 | 157 |
| *KMT2D* | chr12 | 49026171 | 49027332 | 1,161 |
| *KMT2D* | chr12 | 49027792 | 49027940 | 148 |
| *KMT2D* | chr12 | 49027998 | 49028151 | 153 |
| *KMT2D* | chr12 | 49028817 | 49028968 | 151 |
| *KMT2D* | chr12 | 49029050 | 49029246 | 196 |
| *KMT2D* | chr12 | 49029390 | 49029486 | 96 |
| *KMT2D* | chr12 | 49030269 | 49030778 | 509 |
| *KMT2D* | chr12 | 49030882 | 49031043 | 161 |
| *KMT2D* | chr12 | 49031164 | 49033974 | 2,810 |
| *KMT2D* | chr12 | 49034056 | 49034309 | 253 |
| *KMT2D* | chr12 | 49034399 | 49034486 | 87 |
| *KMT2D* | chr12 | 49034571 | 49034676 | 105 |
| *KMT2D* | chr12 | 49034801 | 49034945 | 144 |
| *KMT2D* | chr12 | 49035564 | 49036027 | 463 |
| *KMT2D* | chr12 | 49037114 | 49038999 | 1,885 |
| *KMT2D* | chr12 | 49039211 | 49039368 | 157 |
| *KMT2D* | chr12 | 49039424 | 49039627 | 203 |
| *KMT2D* | chr12 | 49039713 | 49041545 | 1,832 |
| *KMT2D* | chr12 | 49041644 | 49041715 | 71 |
| *KMT2D* | chr12 | 49041906 | 49042000 | 94 |

| Gene | Chromosome | Start | End | Size (bp) |
|---|---|---|---|---|
| *KMT2D* | chr12 | 49042078 | 49042340 | 262 |
| *KMT2D* | chr12 | 49042550 | 49042655 | 105 |
| *KMT2D* | chr12 | 49042730 | 49042888 | 158 |
| *KMT2D* | chr12 | 49043065 | 49043196 | 131 |
| *KMT2D* | chr12 | 49043352 | 49043792 | 440 |
| *KMT2D* | chr12 | 49043857 | 49044008 | 151 |
| *KMT2D* | chr12 | 49044180 | 49044314 | 134 |
| *KMT2D* | chr12 | 49044392 | 49044532 | 140 |
| *KMT2D* | chr12 | 49044733 | 49044975 | 242 |
| *KMT2D* | chr12 | 49045909 | 49045977 | 68 |
| *KMT2D* | chr12 | 49046054 | 49046184 | 130 |
| *KMT2D* | chr12 | 49046249 | 49046434 | 185 |
| *KMT2D* | chr12 | 49046598 | 49046800 | 202 |
| *KMT2D* | chr12 | 49047954 | 49048079 | 125 |
| *KMT2D* | chr12 | 49048648 | 49048779 | 131 |
| *KMT2D* | chr12 | 49049094 | 49049228 | 134 |
| *KMT2D* | chr12 | 49049671 | 49050800 | 1,129 |
| *KMT2D* | chr12 | 49050875 | 49052434 | 1,559 |
| *KMT2D* | chr12 | 49052553 | 49052719 | 166 |
| *KMT2D* | chr12 | 49052904 | 49053082 | 178 |
| *KMT2D* | chr12 | 49053196 | 49053331 | 135 |
| *KMT2D* | chr12 | 49053465 | 49053651 | 186 |
| *KMT2D* | chr12 | 49053967 | 49054150 | 183 |
| *KMT2D* | chr12 | 49054296 | 49054426 | 130 |
| *KMT2D* | chr12 | 49054517 | 49054761 | 244 |
| *KMT2D* | chr12 | 49054889 | 49055036 | 147 |
| *KMT2D* | chr12 | 49055265 | 49055371 | 106 |
| *KMT2D* | chr12 | 49059602 | 49059784 | 182 |
| *MLH1* | chr3 | 36993321 | 36993900 | 579 |
| *MLH1* | chr3 | 36994216 | 36994356 | 140 |
| *MLH1* | chr3 | 36994819 | 36994963 | 144 |
| *MLH1* | chr3 | 36996608 | 36996719 | 111 |
| *MLH1* | chr3 | 36997145 | 36997310 | 165 |
| *MLH1* | chr3 | 37000944 | 37001064 | 120 |
| *MLH1* | chr3 | 37004390 | 37004484 | 94 |
| *MLH1* | chr3 | 37005850 | 37005976 | 126 |
| *MLH1* | chr3 | 37006980 | 37007073 | 93 |

| Gene | Chromosome | Start | End | Size (bp) |
|---|---|---|---|---|
| *MLH1* | chr3 | 37008803 | 37008915 | 112 |
| *MLH1* | chr3 | 37011809 | 37011872 | 63 |
| *MLH1* | chr3 | 37012000 | 37012109 | 109 |
| *MLH1* | chr3 | 37014421 | 37014554 | 133 |
| *MLH1* | chr3 | 37017495 | 37017609 | 114 |
| *MLH1* | chr3 | 37020299 | 37020473 | 174 |
| *MLH1* | chr3 | 37025626 | 37026017 | 391 |
| *MLH1* | chr3 | 37028773 | 37028942 | 169 |
| *MLH1* | chr3 | 37040175 | 37040304 | 129 |
| *MLH1* | chr3 | 37042257 | 37042341 | 84 |
| *MLH1* | chr3 | 37047508 | 37047750 | 242 |
| *MLH1* | chr3 | 37048506 | 37048619 | 113 |
| *MLH1* | chr3 | 37048893 | 37049027 | 134 |
| *MLH1* | chr3 | 37050475 | 37050928 | 453 |
| *STAT3* | chr17 | 42313313 | 42317572 | 4,259 |
| *STAT3* | chr17 | 42322271 | 42322504 | 233 |
| *STAT3* | chr17 | 42322993 | 42323635 | 642 |
| *STAT3* | chr17 | 42324700 | 42324856 | 156 |
| *STAT3* | chr17 | 42324952 | 42325167 | 215 |
| *STAT3* | chr17 | 42326105 | 42326209 | 104 |
| *STAT3* | chr17 | 42329399 | 42329467 | 68 |
| *STAT3* | chr17 | 42329543 | 42329786 | 243 |
| *STAT3* | chr17 | 42331461 | 42331548 | 87 |
| *STAT3* | chr17 | 42333662 | 42333775 | 113 |
| *STAT3* | chr17 | 42333880 | 42334059 | 179 |
| *STAT3* | chr17 | 42337424 | 42337596 | 172 |
| *STAT3* | chr17 | 42337752 | 42337867 | 115 |
| *STAT3* | chr17 | 42338720 | 42338822 | 102 |
| *STAT3* | chr17 | 42339303 | 42339419 | 116 |
| *STAT3* | chr17 | 42345403 | 42345667 | 264 |
| *STAT3* | chr17 | 42346558 | 42346723 | 165 |
| *STAT3* | chr17 | 42348378 | 42348549 | 171 |
| *STAT3* | chr17 | 42374048 | 42374216 | 168 |
| *STAT3* | chr17 | 42386928 | 42388578 | 1,650 |
| *TET2* | chr4 | 105145864 | 105146096 | 232 |
| *TET2* | chr4 | 105146282 | 105146989 | 707 |
| *TET2* | chr4 | 105147446 | 105147809 | 363 |

| Gene | Chromosome | Start | End | Size (bp) |
|---|---|---|---|---|
| *TET2* | chr4 | 105190349 | 105190713 | 364 |
| *TET2* | chr4 | 105202329 | 105202556 | 227 |
| *TET2* | chr4 | 105233886 | 105242781 | 8,895 |
| *TET2* | chr4 | 105242823 | 105242937 | 114 |
| *TET2* | chr4 | 105243559 | 105243788 | 229 |
| *TET2* | chr4 | 105259608 | 105259779 | 171 |
| *TET2* | chr4 | 105261748 | 105261858 | 110 |
| *TET2* | chr4 | 105269599 | 105269757 | 158 |
| *TET2* | chr4 | 105272553 | 105272928 | 375 |
| *TET2* | chr4 | 105275037 | 105279826 | 4,789 |
| *TP53* | chr17 | 7661768 | 7662024 | 256 |
| *TP53* | chr17 | 7665405 | 7665541 | 136 |
| *TP53* | chr17 | 7666075 | 7666254 | 179 |
| *TP53* | chr17 | 7666891 | 7667435 | 544 |
| *TP53* | chr17 | 7668391 | 7669700 | 1,309 |
| *TP53* | chr17 | 7670598 | 7670725 | 127 |
| *TP53* | chr17 | 7673196 | 7673349 | 153 |
| *TP53* | chr17 | 7673524 | 7673618 | 94 |
| *TP53* | chr17 | 7673690 | 7673847 | 157 |
| *TP53* | chr17 | 7674170 | 7674300 | 130 |
| *TP53* | chr17 | 7674515 | 7674981 | 466 |
| *TP53* | chr17 | 7675042 | 7675503 | 461 |
| *TP53* | chr17 | 7675983 | 7676282 | 299 |
| *TP53* | chr17 | 7676371 | 7676632 | 261 |
| *TP53* | chr17 | 7677314 | 7677444 | 130 |
| *TP53* | chr17 | 7685249 | 7686381 | 1,132 |
| *TP53* | chr17 | 7687366 | 7687560 | 194 |

Target panel designed using Agilent SureSelect SureDesign (Design ID: 3497811). Coordinates based on human reference genome hg38 (GRCh38). Total panel size: 168.5 kb across 284 target regions.

**Supplementary Table 3. Artificial mosaic spike-in sensitivity assay**

| Spike-in VAF | MS1 Mutect2 | MS2 Mutect2 | Duplex Mutect2 | Duplex Read-level | True Sensitivity |
|---|---|---|---|---|---|
| 1.00% | 100% | 100% | 100% | 100% | 100% |
| 0.50% | 93.2% | 79.5% | 65.9% | 88.6% | 88.6% |
| 0.25% | 81.8% | 54.5% | 22.7% | 75.0% | 70.5% |
| 0.125% | 41.9% | 9.3% | 7.0% | 34.9% | 18.6% |

Sensitivity was assessed by spiking G51D brain-derived DNA into a control sample at defined dilutions (see Methods). Source DNA carried 43 heterozygous germline SNPs and one homozygous SNP (germline VAFs ~0.5 and ~1.0 respectively); dilutions of 1%, 0.5%, and 0.25% produced expected spike-in VAFs of ~0.5%, ~0.25%, and ~0.125% for heterozygous SNPs, and ~1.0%, ~0.5%, and ~0.25% for the homozygous SNP. Total variants assessed: 1.00% (n=1), 0.50% (n=44), 0.25% (n=44), 0.125% (n=43). MS1 = Hybrid-MS1 consensus; MS2 = Hybrid-MS2 consensus; Duplex = full duplex consensus (both DNA strands required). Mutect2 columns: proportion of spike-in variants actively called by Mutect2 at the respective consensus stringency. Duplex Read-level: proportion of spike-in variants with at least one supporting alt read in the Duplex consensus BAM. True Sensitivity: requires both (i) Mutect2 calling in at least one stringency level, AND (ii) at least one supporting alt read in the Duplex BAM.

**Supplementary Table 4. Somatic variant filtering parameters and thresholds**

| Filter | Parameter | Rationale | Threshold |
|---|---|---|---|
| Base Quality | MBQ | Low-quality bases have higher sequencing error probability | ≥ Q30 |
| Allele Fraction | VAF | Somatic variants expected at low VAF; high VAF suggests germline | ≤ 0.3 |
| Strand Bias | SOR | Excessive strand bias indicates strand-specific sequencing artifacts | ≤ 3 |
| Mapping Quality | MMQ | Low mapping quality indicates ambiguous read alignment in repetitive regions | ≥ 40 |
| Clustered Variants | ECNT | Variant clusters suggest localized alignment errors | ≤ 2 |
| Read Position | MPOS | Sequencing error rates and library artifacts increase toward read ends | ≥ 5 |
| Recurrent Artifact Exclusion | Alt Reads | Recurrent calls across many samples suggests common sequencing error or contamination | Excluded candidates in multiple individuals |
| Repetitive Regions | RepeatMasker (indels only) | Indel calls in repetitive sequences prone to alignment artifacts | Excluded |
| Manual Inspection | IGV | Final validation to exclude technical artifacts and confirm variant authenticity | Required |

MBQ = Median Base Quality; VAF = variant allele fraction; SOR = symmetric odds ratio; MMQ = median mapping quality; ECNT = event count within 10 bp; MPOS = median position from end of read. All quality metrics (rows 1–6) derived from Mutect2 (GATK v4.6.1.0) VCF annotations. RepeatMasker filtering applied to indels only. Manual IGV inspection performed as final validation step.

**Supplementary Table 5. CH mutation coordinates queried by targeted duplex mpileup and cross-referenced against Mutect2 callset**

| Gene | Codon / Mutation | Co-ordinates | Mutation type | Notes | Detected in this study | Reference(s) |
|---|---|---|---|---|---|---|
| *ASXL1* | W522X | chr20:32,434,460 | Stop-gain | Stop gain; confirmed CH whole blood (1.72% VAF in one carrier) | — | Wiegand 2026 |
| *CBL* | C404 | chr11:119,278,281 | Missense | C404Y recurrent mutation in blood cancer (10%); RING finger domain | — | Lim 2024 |
| *CBL* | R420 | chr11:119,278,541 | Missense | R420Q recurrent mutation in blood cancer (9%); abolishes CBL E3 ubiquitin ligase activity | — | Lim 2024 |
| *DNMT3A* | R214H | chr2:25,247,076 | Missense | Confirmed CH in multipotent progenitors (24.3% VAF in one carrier) | — | Wiegand 2026 |
| *DNMT3A* | R577W | chr2:25,240,439 | Missense | Confirmed CH whole blood; commonly found in healthy individuals | — | Wiegand 2026 |
| *DNMT3A* | R635W/Q | chr2:25,243,931 | Missense | Commonly found in CH in healthy individuals | — | Bick 2020; Vlasschaert 2022 |
| *DNMT3A* | K674R | chr2:25,236,937 | Missense | Confirmed CH in multipotent progenitors (2.4% VAF in one carrier) | — | Wiegand 2026 |
| *DNMT3A* | D702 | chr2:25,240,708 | Missense | Methyltransferase domain; functionally pLoF; multiple distinct substitutions (D702E/V/Y/H/N) | DNMT3A_SNV2 (SND-04); Mutect2 | Bick 2020; Niroula 2021; Vlasschaert 2022 |
| *DNMT3A* | R688C | chr2:25,241,582 | Missense | Commonly found in CH in healthy individuals | DNMT3A_SNV3 (SND-09); mpileup | Bick 2020; Vlasschaert 2022 |
| *DNMT3A* | R882H/C/S | chr2:25,234,373/ 25,234,374 | Missense | R882H: most prevalent CH hotspot; first described in AML and confirmed in healthy aging; dominant negative mechanism reducing methyltransferase activity by ~80%. R882C and R882S less common. | — | Ley 2010; McKerrell 2015; Bick 2020; Niroula 2021; Vlasschaert 2022 |
| *DNMT3A* | A884W/V | chr2:25,234,367 | Missense | Adjacent to R882 hotspot; commonly found in CH | — | Bick 2020; Niroula 2021; Vlasschaert 2022 |
| *KMT2D* | L1055X | chr12:49,050,424 | Stop-gain | Confirmed in multipotent progenitors in lymphoma | — | Wiegand 2026 |
| *KMT2D* | R5432Q | chr12:49,022,633 | Missense | L-CHIP driver; confirmed in lymphoma progenitor cells | — | Niroula 2021; Wiegand 2026 |

| Gene | Codon / Mutation | Co-ordinates | Mutation type | Notes | Detected in this study | Reference(s) |
|---|---|---|---|---|---|---|
| *STAT3* | S614N/R | chr17:42,323,051 | Missense | Confirmed in leukemia; named L-CHIP driver | — | Niroula 2021; Olson 2021 |
| *STAT3* | E616G/K | chr17:42,323,045 | Missense | STAT3 activation domain hotspot; named L-CHIP driver | — | Niroula 2021 |
| *STAT3* | G618R | chr17:42,323,040 | Missense | STAT3 activation domain; GOF functionally validated; confirmed in leukemia | — | Olson 2021 |
| *STAT3* | Y640F | chr17:42,322,464 | Missense | Most common STAT3-CH mutation; cytokine-independent GOF activation confirmed in vitro | — | Koskela 2012; Niroula 2021; Olson 2021; Raess 2017 |
| *STAT3* | N647I | chr17:42,322,443 | Missense | 4th most common in leukemia (3/77, 4%) | — | Koskela 2012; Olson 2021 |
| *STAT3* | K658R/M | chr17:42,322,410 | Missense | STAT3 activation domain hotspot; L-CHIP criterion | — | Niroula 2021; Koskela 2012; Olson 2021 |
| *STAT3* | I659L | chr17:42,322,408 | Missense | STAT3 activation domain; confirmed in leukemia; Sanger validated | — | Olson 2021 |
| *STAT3* | D661V/Y | chr17:42,322,401 | Missense | Joint 2nd most common in leukemia (7/77 each, 9%); GOF confirmed in vitro | — | Koskela 2012; Niroula 2021; Olson 2021 |
| *TET2* | Y437X | chr4:105,235,253 | Stop-gain | Confirmed CH whole blood (1.17% VAF in one carrier) | — | Wiegand 2026 |
| *TET2* | R550X | chr4:105,235,590 | Stop-gain | High-confidence pLoF; confirmed in leukemia (40% VAF) | — | Olson 2021 |
| *TET2* | Q720X | chr4:105,236,100 | Stop-gain | pLoF; confirmed in leukemia (14% VAF) | — | Olson 2021 |
| *TET2* | E1144K | chr4:105,241,359 | Missense | TET2 catalytic domain; confirmed in leukemia (40% VAF) | — | Raess 2017 |
| *TET2* | G1184D | chr4:105,242,884 | Missense | TET2 catalytic domain; pLoF; confirmed in leukemia (9% VAF) | — | Olson 2021 |
| *TET2* | E1268X | chr4:105,243,777 | Stop-gain | TET2 catalytic domain; pLoF; confirmed in leukemia (43% VAF); Sanger validated | — | Olson 2021 |
| *TET2* | R1452X | chr4:105,272,735 | Stop-gain | TET2 catalytic domain; pLoF; confirmed in leukemia (22% VAF) | — | Olson 2021 |

| Gene | Codon / Mutation | Co-ordinates | Mutation type | Notes | Detected in this study | Reference(s) |
|---|---|---|---|---|---|---|
| *TET2* | R1465X | chr4:105,272,774 | Stop-gain | TET2 catalytic domain; confirmed CH whole blood and progenitor fractions (5.4% VAF) | — | Wiegand 2026 |
| *TET2* | A1512V | chr4:105,272,916 | Missense | TET2 catalytic domain; confirmed CH whole blood (2.5% VAF) | — | Wiegand 2026 |
| *TET2* | R1516X | chr4:105,275,056 | Stop-gain | TET2 catalytic domain; confirmed CH whole blood (10.8% VAF) | — | Wiegand 2026 |
| *TET2* | G1860E | chr4:105,276,089 | Missense | TET2 catalytic domain; confirmed in leukemia (25% VAF) | — | Olson 2021 |
| *TET2* | L1872R | chr4:105,276,125 | Missense | TET2 catalytic domain; confirmed in leukemia (12% VAF); Sanger validated | — | Olson 2021 |
| *TET2* | V1900I | chr4:105,276,208 | Missense | TET2 catalytic domain; confirmed in CH (4.6% VAF) | — | Wiegand 2026 |
| *TP53* | R273H/C | chr17:7,673,802 | Missense | Most common TP53-CHIP codon; 80 total carriers; median VAF 3.4–4.2%; DNA contact mutant | — | Usui 2025; Bick 2020; Niroula 2021; Vlasschaert 2022 |
| *TP53* | Y220C | chr17:7,674,872 | Missense | 56 carriers; median VAF 3.8%; structural mutant | — | Usui 2025 |
| *TP53* | R248Q/W | chr17:7,674,220 | Missense | 71 total carriers; median VAF 3.9–5.4%; DNA contact mutant | TP53_SNV2 (SND-06); mpileup | Usui 2025; Bick 2020; Niroula 2021; Vlasschaert 2022 |
| *TP53* | R175H | chr17:7,675,088 | Missense | 34 carriers; median VAF 4.0%; structural mutant | — | Usui 2025; Bick 2020; Niroula 2021; Vlasschaert 2022 |
| *TP53* | G245S/D | chr17:7,674,230 | Missense | 35 carriers; median VAF 3.7–4.9%; DNA contact mutant | — | Usui 2025 |
| *TP53* | R282W | chr17:7,673,776 | Missense | 21 carriers; median VAF 4.1%; structural mutant | — | Usui 2025 |
| *TP53* | C238Y | chr17:7,674,250 | Missense | 21 carriers; median VAF 4.5%; max VAF 32.2% | — | Usui 2025 |
| *TP53* | V216M | chr17:7,674,885 | Missense | 17 carriers; median VAF 3.6% | — | Usui 2025 |
| *TP53* | V272M | chr17:7,673,806 | Missense | 16 carriers; median VAF 3.5%; max VAF 21.4% | — | Usui 2025 |

| Gene | Codon / Mutation | Co-ordinates | Mutation type | Notes | Detected in this study | Reference(s) |
|---|---|---|---|---|---|---|
| *TP53* | H179R/L | chr17:7,675,076 | Missense | 18 carriers; median VAF 5.8–12.5%; DNA contact mutant | — | Usui 2025 |
| *TP53* | C275Y | chr17:7,673,796 | Missense | 13 carriers; median VAF 4.2% | — | Usui 2025 |
| *TP53* | V173M | chr17:7,675,095 | Missense | 12 carriers; median VAF 6.0%; max VAF 30.7% | — | Usui 2025 |
| *TP53* | K132R | chr17:7,675,217 | Missense | 12 carriers; median VAF 3.9%; max VAF 25.3% | — | Usui 2025 |
| *TP53* | V143M | chr17:7,675,185 | Missense | 12 carriers; median VAF 3.6%; max VAF 20.2% | — | Usui 2025 |
| *TP53* | S241F | chr17:7,674,241 | Missense | 11 carriers; median VAF 5.6%; max VAF 34.6% | — | Usui 2025 |
| *TP53* | C277Y | chr17:7,673,790 | Missense | 11 carriers; median VAF 6.1%; max VAF 32.0% | — | Usui 2025 |
| *TP53* | R158H | chr17:7,675,139 | Missense | 11 carriers; median VAF 3.2%; max VAF 24.3% | — | Usui 2025 |
| *TP53* | M237I | chr17:7,674,252 | Missense | 11 carriers; median VAF 3.0% | — | Usui 2025 |
| *TP53* | H193R | chr17:7,674,953 | Missense | 10 carriers; median VAF 4.1% | — | Usui 2025 |
| *TP53* | Y234C | chr17:7,674,262 | Missense | 10 carriers; median VAF 3.4% | — | Usui 2025 |
| *TP53* | G244D | chr17:7,674,232 | Missense | 6 carriers; median VAF 13.1%; max VAF 27.8% | — | Usui 2025 |
| *TP53* | c.376-1G>A (splice) | chr17:7,675,993 | Splice-acceptor | 7 carriers; median VAF 11.2%; max VAF 23.3% | — | Usui 2025 |

This table was compiled by systematic review of published clonal hematopoiesis (CH) cohort studies and targeted sequencing datasets. For each position, inclusion required direct observation of a mutation at the specific residue in at least one published carrier. All genomic coordinates are GRCh38/hg38, verified against dbSNP/VarSome, and confirmed within our capture panel. 'Detected in this study' indicates residues at which a variant was identified either by Mutect2 variant calling or targeted mpileup of duplex BAMs; detection method specified per entry. Abbreviations: M-CHIP, myeloid clonal hematopoiesis of indeterminate potential; L-CHIP, lymphoid clonal hematopoiesis of indeterminate potential; GOF, gain-of-function; pLoF, predicted loss-of-function. Bick 2020 = Bick et al., Nature 2020; Niroula 2021 = Niroula et al., Nat Med 2021; Vlasschaert 2022 = Vlasschaert et al., Blood 2022; Usui 2025 = Usui et al., Blood Cancer Discov 2025; Wiegand 2026 = Wiegand et al., Blood 2026; Lim 2024 = Lim et al., PLoS One 2024; Koskela 2012 = Koskela et al., NEJM 2012; Olson 2021 = Olson et al., Blood 2021; Raess 2017 = Raess et al., Am J Hematol 2017; Ley 2010 = Ley et al., N Engl J Med 2010; McKerrell 2015 = McKerrell et al., Cell Rep 2015.

**Supplementary Table 6. Sequencing coverage statistics across error correction stringencies**

| Sample ID | Region | Raw (×) | MS1 (×) | MS2 (×) | Duplex (×) |
|---|---|---|---|---|---|
| OPCA-01 | CER | 14,398 | 8,769 | 2,687 | 1,241 |
| OPCA-01 | CC | 8,566 | 5,560 | 1,546 | 693 |
| OPCA-01 | PUT | 9,226 | 6,210 | 1,626 | 693 |
| OPCA-02 | CER | 10,794 | 6,811 | 1,999 | 857 |
| OPCA-02 | PUT | 11,370 | 6,679 | 2,180 | 990 |
| OPCA-03 | CER | 14,224 | 8,536 | 2,814 | 1,046 |
| OPCA-03 | CC | 14,355 | 8,245 | 2,829 | 1,179 |
| OPCA-03 | PUT | 12,905 | 8,840 | 2,201 | 1,039 |
| OPCA-05 | CER | 9,528 | 6,266 | 1,740 | 692 |
| OPCA-05 | CC | 9,699 | 7,026 | 1,561 | 684 |
| OPCA-05 | PUT | 14,007 | 8,026 | 2,895 | 921 |
| OPCA-06 | CER | 12,984 | 7,963 | 2,368 | 1,186 |
| OPCA-06 | CC | 9,993 | 7,011 | 1,653 | 751 |
| OPCA-06 | PUT | 11,559 | 7,794 | 1,979 | 921 |
| OPCA-07 | CER | 14,854 | 9,209 | 2,746 | 1,249 |
| OPCA-07 | CC | 11,558 | 7,760 | 1,999 | 945 |
| OPCA-07 | PUT | 13,260 | 6,938 | 2,506 | 1,296 |
| OPCA-08 | CER | 12,848 | 7,920 | 2,382 | 1,092 |
| OPCA-08 | CC | 13,931 | 8,194 | 2,664 | 1,192 |
| OPCA-08 | PUT | 13,480 | 8,511 | 2,432 | 1,198 |
| OPCA-09 | CER | 11,534 | 7,229 | 2,125 | 952 |
| OPCA-09 | CC | 12,916 | 7,736 | 2,442 | 1,114 |
| OPCA-09 | PUT | 10,580 | 6,232 | 1,981 | 959 |
| OPCA-11 | CER | 11,728 | 7,400 | 2,111 | 1,043 |
| OPCA-11 | CC | 10,790 | 6,765 | 2,034 | 834 |
| OPCA-11 | PUT | 11,876 | 7,631 | 2,165 | 947 |
| OPCA-12 | CER | 9,567 | 5,359 | 1,766 | 938 |
| OPCA-12 | CC | 11,125 | 7,260 | 2,035 | 807 |
| OPCA-12 | PUT | 12,376 | 8,133 | 2,155 | 1,075 |
| SND-01 | CER | 15,428 | 7,618 | 3,050 | 1,362 |
| SND-01 | CC | 10,600 | 6,625 | 1,927 | 945 |
| SND-01 | PUT | 10,985 | 7,636 | 1,821 | 850 |
| SND-04 | CER | 11,837 | 7,481 | 2,212 | 936 |

| Sample ID | Region | Raw (×) | MS1 (×) | MS2 (×) | Duplex (×) |
|---|---|---|---|---|---|
| SND-04 | CC | 9,018 | 5,692 | 1,699 | 691 |
| SND-04 | PUT | 10,709 | 7,072 | 1,916 | 850 |
| SND-05 | CER | 10,695 | 6,098 | 2,090 | 908 |
| SND-05 | CC | 9,490 | 6,041 | 1,729 | 796 |
| SND-05 | PUT | 12,171 | 7,050 | 2,379 | 988 |
| SND-06 | CER | 11,542 | 7,025 | 2,165 | 991 |
| SND-06 | CC | 10,387 | 6,146 | 2,009 | 814 |
| SND-06 | PUT | 11,427 | 6,650 | 2,263 | 855 |
| SND-07 | CER | 8,617 | 5,692 | 1,517 | 691 |
| SND-07 | CC | 10,203 | 6,932 | 1,743 | 821 |
| SND-07 | PUT | 10,520 | 7,323 | 1,746 | 812 |
| SND-08 | CER | 13,117 | — | 2,639 | 974 |
| SND-08 | CC | 14,496 | 9,255 | 2,608 | 1,272 |
| SND-08 | PUT | 14,898 | 7,533 | 2,850 | 1,398 |
| SND-09 | CER | 11,779 | 6,615 | 2,317 | 992 |
| SND-09 | CC | 12,847 | 7,255 | 2,532 | 1,034 |
| SND-09 | PUT | 10,424 | 6,366 | 1,973 | 858 |
| SND-13 | CER | 10,577 | 6,464 | 2,001 | 867 |
| SND-13 | CC | 11,518 | 6,077 | 2,434 | 761 |
| SND-13 | PUT | 6,090 | 4,637 | 885 | 418 |
| SND-15 | CER | 10,152 | 6,676 | 1,776 | 881 |
| SND-15 | CC | 8,817 | 5,980 | 1,510 | 718 |
| SND-15 | PUT | 10,974 | 7,155 | 1,972 | 885 |
| SND-16 | CER | 11,336 | 7,333 | 2,069 | 876 |
| SND-16 | CC | 12,946 | 7,715 | 2,449 | 1,110 |
| SND-16 | PUT | 11,697 | 7,697 | 2,074 | 949 |
| Control-02 | CER | 11,982 | 7,518 | 2,227 | 974 |
| Control-02 | CC | 10,602 | 6,275 | 2,074 | 828 |
| Control-02 | PUT | 11,248 | 6,743 | 2,164 | 874 |
| Control-03 | CER | 12,836 | 8,582 | 2,289 | 953 |
| Control-03 | CC | 12,240 | 8,382 | 2,056 | 1,016 |
| Control-03 | PUT | 11,883 | 7,589 | 2,165 | 953 |
| Control-04 | CER | 13,475 | 8,491 | 2,411 | 1,222 |
| Control-04 | CC | 11,659 | 7,357 | 2,131 | 980 |

| Sample ID | Region | Raw (×) | MS1 (×) | MS2 (×) | Duplex (×) |
|---|---|---|---|---|---|
| Control-04 | PUT | 11,510 | 7,386 | 2,037 | 1,039 |
| Control-05 | CER | 14,275 | — | 2,781 | 1,178 |
| Control-05 | CC | 13,182 | 7,399 | 2,529 | 1,178 |
| Control-05 | PUT | 12,339 | 7,353 | 2,307 | 1,103 |
| Control-06 | CER | 13,757 | 7,698 | 2,647 | 1,220 |
| Control-06 | CC | 10,225 | 7,092 | 1,680 | 883 |
| Control-06 | PUT | 14,492 | 7,744 | 2,849 | 1,227 |
| Control-07 | CER | 10,510 | 6,889 | 1,848 | 911 |
| Control-07 | CC | 11,574 | 6,623 | 2,185 | 1,123 |
| Control-07 | PUT | 11,274 | 6,826 | 2,176 | 891 |
| Control-08 | CER | 9,142 | 5,957 | 1,632 | 750 |
| Control-08 | CC | 10,656 | 6,837 | 1,955 | 844 |
| Control-08 | PUT | 9,404 | 6,323 | 1,642 | 745 |
| Control-11 | CER | 12,746 | 8,447 | 2,313 | 919 |
| Control-11 | CC | 14,086 | 8,574 | 2,817 | 927 |
| Control-11 | PUT | 13,837 | 8,542 | 2,663 | 1,021 |
| Control-14 | CER | 12,489 | 7,677 | 2,333 | 1,047 |
| Control-14 | CC | 14,762 | 8,854 | 2,840 | 1,194 |
| Control-14 | PUT | 11,267 | 7,087 | 2,060 | 952 |

Coverage depth (mean ×) for all samples across four consensus stringency levels: Raw (unprocessed BAM), MS1 (Hybrid-MS1 consensus), MS2 (Hybrid-MS2 consensus), and Duplex (full duplex consensus, both DNA strands). Target panel size: 168.5 kb. CER = cerebellar white matter; CC = cingulate cortex; PUT = putamen. Mann-Whitney U tests showed no significant differences between MSA and control groups for any stringency level across all brain regions (all $p>0.05$). Two samples had missing MS1 data (SND-08 cerebellum, Control-05 cerebellum).

### Supplementary Table 7. Complete somatic mutation callset

| Variant ID | Mutation | Consequence | Protein change | CADD-Phred | Case ID | CER | CC | PUT |
|---|---|---|---|---|---|---|---|---|
| *ATRX*_DEL1 | chrX:g.77785985_77785987del | Inframe deletion | p.Met6del | 17.95 | SND-04 | — | 0.68 | — |
| *BCR*_SNV2 | chr22:g.23316722C>T | 3' UTR | — | 0.20 | OPCA-09 | 7.58 | 7.25 | 6.75 |
| *BCR*_SNV3 | chr22:g.23316738G>C | 3' UTR | — | 1.08 | OPCA-09 | 6.89 | 6.46 | 6.03 |
| *BCR*_DEL1 | chr22:g.23260930_23260933del | Intronic | — | 2.35 | Control-05 | 0.34 | — | — |
| *BCR*_DEL2 | chr22:g.23261424_23261427del | — | — | 21.70 | OPCA-07 | 0.07 | — | — |
| *BCR*_DEL3 | chr22:g.23268535_23268538del | Intronic | — | 3.48 | OPCA-01 | 0.27 | — | — |
| *BCR*_DEL4 | chr22:g.23290839_23290840del | Intronic | — | 1.57 | Control-04 | 0.52 | — | — |
| *BCR*_DEL5 | chr22:g.23316590del | 3' UTR | — | 2.91 | Control-03 | — | 8.30 | 11.10 |
| *CBL*_SNV2 | chr11:g.119278528T>C | Missense | p.Cys416Arg | 29.90 | Control-05 | 1.47 | — | — |
| *DNMT3A*_SNV1 | chr2:g.25240652T>A | Stop gained | p.Lys721Ter | 42.00 | Control-08 | 1.02 | — | — |
| *DNMT3A*_SNV2 | chr2:g.25240708T>C | Missense | p.Asp702Gly | 30.00 | SND-04 | 0.93 | 2.17 | 0.88 |
| *DNMT3A*_SNV3 | chr2:g.25241582C>T | Missense | p.Arg688Cys | 31.00 | SND-09 | — | 0.21 | 0.15 |
| *DNMT3A*_DEL1 | chr2:g.25247067_25247081del | Inframe deletion | p.Met364_Ala368del | 22.50 | SND-08 | 0.38 | 0.35 | 0.54 |
| *KCTD7*_SNV1 | chr7:g.66633386T>C | Missense | p.Tyr86His | 19.81 | Control-08 | — | 0.24 | 0.36 |
| *KCTD7*_SNV2 | chr7:g.66638677T>A | Intronic | — | 1.10 | Control-05 | — | — | 0.10 |
| *KCTD7*_DEL1 | chr7:g.66629001_66629011del | 5' UTR | — | 18.04 | SND-04 | — | 0.45 | — |
| *KCTD7*_DEL2 | chr7:g.66629071_66629073del | Inframe deletion | p.Val4del | 20.70 | OPCA-03 | — | 0.08 | 0.24 |
| *KMT2D*_SNV1 | chr12:g.49022275C>T | Intronic | — | 22.50 | OPCA-06 | — | 0.36 | — |
| *KMT2D*_SNV3 | chr12:g.49034200C>T | Missense | p.Arg3536His | 27.60 | SND-01 | — | 0.40 | 0.34 |
| *KMT2D*_DEL1 | chr12:g.49024674_49024677del | Frameshift | p.Leu5318SerfsTer14 | 36.00 | SND-15 | — | 0.40 | — |
| *KMT2D*_DEL2 | chr12:g.49033837_49033839del | Inframe deletion | p.Gln3623del | 18.21 | OPCA-05 | — | — | 0.67 |
| *KMT2D*_INS | chr12:g.49042006_49042007insGGGT | Intronic | — | 1.54 | SND-15 | — | — | 0.26 |
| *MLH1*_DEL1 | chr3:g.37025890_37025893del | Frameshift | p.Leu432AsnfsTer58 | 34.00 | Control-08 | 0.35 | — | — |
| *SNCA*_SNV1 | chr4:g.89725768A>T | 3' UTR | — | 3.98 | Control-08 | — | 0.60 | 0.11 |
| *SNCA*_SNV2 | chr4:g.89726714T>C | Intronic | — | 4.36 | Control-08 | — | 0.35 | 0.04 |
| *SNCA*_SNV3 | chr4:g.89822814A>G | Intronic | — | 0.65 | Control-06 | 0.06 | — | 0.03 |
| *SNCA*_SNV4 | chr4:g.89836785T>C | Intronic | — | 3.69 | OPCA-06 | — | 0.61 | 0.34 |
| *SNCA*_SNV5 | chr4:g.89838202T>C | Upstream | — | 7.91 | SND-01 | 0.24 | 0.37 | 0.77 |

| Variant ID | Mutation | Consequence | Protein change | CADD-Phred | Case ID | CER | CC | PUT |
|---|---|---|---|---|---|---|---|---|
| *SNCA*_DEL1 | chr4:g.89720616_89720618del | Intronic | — | 6.06 | Control-05 | — | 0.80 | — |
| *SNCA*_DEL2 | chr4:g.89801500_89801504del | Intronic | — | 0.83 | Control-14 | — | 0.32 | — |
| *STAT3*_DEL1 | chr17:g.42334057_42334061del | Intronic | — | 7.87 | SND-15 | — | — | 0.48 |
| *TET2*_SNV1 | chr4:g.105275662G>T | Missense | p.Val1718Leu | 1.06 | OPCA-01 | 0.02 | 0.29 | 0.05 |
| *TP53*_SNV2 | chr17:g.7674221G>A | Missense | p.Arg248Trp | 29.20 | SND-06 | 0.39 | 0.39 | — |
| *TP53*_DEL1 | chr17:g.7677410_7677411del | 5' UTR | — | 1.53 | OPCA-12 | 0.44 | — | — |

All somatic mutations detected across MSA (OPCA and SND) brains and controls. Variant IDs match those referenced in the main text. Mutation given in HGVS nomenclature; genomic coordinates based on human reference genome hg38 (GRCh38). Consequence and protein change annotated using OpenCRAVAT; '—' indicates no protein change (non-coding). CADD: Combined Annotation Dependent Depletion score (phred-scaled). Cerebellum (CER), Cingulate Cortex (CC), and Putamen (PUT) columns show VAF (%); '—' indicates variant not detected in that region.
